# Genome-wide analysis of binge-eating disorder identifies the first three risk loci and implicates iron metabolism

**DOI:** 10.1101/2022.04.28.22274437

**Authors:** David Burstein, Trevor Griffen, Karen Therrien, Jaroslav Bendl, Sanan Venkatesh, Pengfei Dong, Amirhossein Modabbernia, Biao Zeng, Deepika Mathur, Gabriel Hoffman, Robyn Sysko, Tom Hildebrandt, Georgios Voloudakis, Panos Roussos

## Abstract

Binge-eating disorder (BED) is the most common eating disorder yet its genetic architecture remains largely unknown. Studying BED is challenging because it is often comorbid with obesity, a common and highly polygenic trait, and it is underdiagnosed in biobank datasets. To address this limitation, we apply a supervised machine learning approach to estimate the probability of each individual having BED based on electronic medical records from the Million Veteran Program. We perform a genome-wide association study on individuals of African (*n* = 77,574) and European (*n* = 285,138) ancestry while controlling for body mass index to identify three independent loci near the *HFE, MCHR2* and *LRP11* genes, which are reproducible across three independent cohorts. We identify genetic association between BED and several neuropsychiatric traits and implicate iron metabolism in the pathophysiology of BED. Overall, our findings provide insights into the genetics underlying BED and suggest directions for future translational research.

---

BED is a common, heritable (41-57%) psychiatric disorder^1,2^ with lifetime prevalence estimated between 0.9 and 3%^3,4^. BED was a provisional diagnosis until the publication of the DSM 5 in 2014 when sufficient evidence was available to conclude that BED can be reliably discriminated from other eating disorders^5^, including anorexia nervosa (AN) or bulimia nervosa (BN). Because BED was only recently added to the most widely used diagnostic classification systems, a sufficiently powered case-control cohort has not yet been recruited^6^ and few subjects can be identified with traditional approaches to electronic medical record (EMR)-driven genetic studies in existing genotyped cohorts. Thus, identification of genetic risk variants for BED has lagged behind similar efforts for other neuropsychiatric disorders. As a result, our understanding of the biological processes underlying the pathophysiology of BED is limited and the development of genetically based, targeted treatments for BED remains out of reach.

BED is epidemiologically associated with obesity, metabolic dysfunction, multiple psychiatric disorders and low overall well-being^4,7^. The heritability of BED is enhanced when adjusting for its association with obesity^1,2^. BED patients have a higher genetic liability for obesity when compared to healthy controls and those with other eating disorders^8^. While research distinguishing BED from obesity is limited, individuals with BED consistently consume more calories in laboratory meals than obese individuals without BED and demonstrate higher levels of psychiatric comorbidity, eating disorder psychopathology, distress and quality of life impairment^5,9,10^. Frequent binge eating is associated with poor metabolic outcomes after adjusting for body mass index (BMI)^11,12^ and those with BED are resistant to sustained weight-loss after interventions with known efficacy^4,13^. Successful treatment of BED does not necessarily result in metabolic improvement or significant weight change^13,14^ and approximately one third of individuals with current or recent BED do not have obesity^3^. Overall, BED is a serious disorder with psychiatric and metabolic components that is epidemiologically distinct from obesity yet shares some underlying genetic influences.

Towards elucidating the genetic architecture of BED, we leveraged the Million Veteran Program (MVP)^15^ to develop and validate a supervised machine-learning algorithm based on clinically diagnosed BED cases to estimate each individual’s likelihood of having BED. By performing a bi-ancestry genome-wide association study (GWAS), we identified three genetic loci that are significantly associated with BED independent of BMI and we implicated a fourth gene through MAGMA gene analysis^16^. We validated our approach by performing polygenic risk score (PRS) analysis in three independent cohorts: the Adolescent Brain and Cognitive Development Study (ABCD)^17^, the Philadelphia Neurodevelopmental Cohort (PNC)^18^ and the UK Biobank (UKBB)^19^. By performing genetic correlation analysis, we found that BED has considerable genetic overlap with several other neuropsychiatric phenotypes, including depression, bipolar disorder and attention-deficit/hyperactivity disorder. Finally, using gene-based and partitioned heritability analyses, we implicate iron metabolism in the pathophysiology of BED.

## Computational Phenotyping Approach

There are significant challenges in identifying individuals with BED using EMR: BED was not fully recognized by the American Psychiatric Association until 2014^20^, BED remains underdiagnosed^21,22^ and the most prevalent disease classification system, the International Classification of Diseases, Ninth Revision, Clinical Modification (ICD-9-CM) does not have a BED-specific diagnostic code. Consequently, in large, longstanding EMRs such as the Veterans Health Administration’s MVP where diagnoses were coded in ICD-9-CM until 2015, most individuals with BED are likely either undiagnosed or diagnosed using codes corresponding to other or unspecified eating disorders (ambiguous codes). Using single ICD codes, we investigated the prevalence of eating disorder diagnoses (*n* = 4,266) within the MVP and found that BED (*n* = 851) was underrepresented relative to BN (*n* = 876) even though BED has a higher prevalence^21^ (Supplementary Table 1). After excluding those who had not been genotyped, there were less than 500 individuals with BED for any given genetic ancestry.

To identify individuals in the MVP with a high likelihood of having BED and allow for the performance of a well-powered GWAS, we developed a machine learning approach reliant upon individuals clinically diagnosed with BED. We first constructed a list of subjects reliably diagnosed with BED (*n* = 822) and a list of controls who did not have any eating disorder diagnoses (*n* = 766,705; Supplementary Table 2; Methods). To calculate a BED-score for each subject in the MVP, we built a LASSO logistic regression model across our BED plus controls cohort (*n* = 767,527). Our hypothesis-free model generated multiple predictors of BED (Top predictors: Fig. 1a; full list: Supplementary Table 3), many of which have known associations with BED. Our model selected multiple medications found to be effective in treating BED in a randomized, controlled trial: lisdexamfetamine, escitalopram, fluoxetine, naltrexone and atomoxetine^23^. The model selected numerous medications used for weight loss and/or treatment of diabetes (bupropion, lorcaserin, phentermine, liraglutide and dulaglutide)^24^ and equipment used in glucose monitoring (glucose sensors). Antipsychotic medications associated with weight gain and diabetes (quetiapine and risperidone)^25^ were selected as negative predictors of BED. The model incorporated broad categories of mood, anxiety and endocrine disorders, all associated with BED^7^. The model also captured predictors related to reproductive organs and sex hormones, gastrointestinal distress, a moisturizer, antivenins and antitoxins, other medications, BMI, female sex, white race and other demographic, diagnostic and EMR factors.

**Fig. 1:**
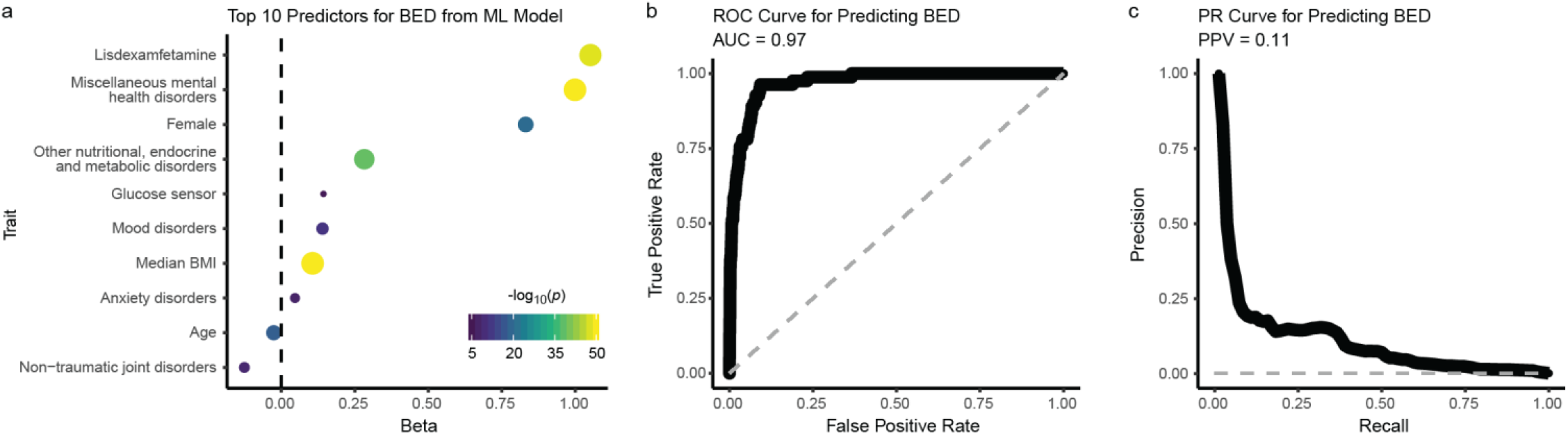
Machine learning model to predict BED within the MVP. **a**, Top 10 predictors from the machine learning lasso logistic regression model for predicting BED (*y* axis). The strength of the statistical association is represented by on the *x* axis as beta and in the size and color of the data points, corresponding to the negative log_10_ of uncorrected two-sided *P* value (-log_10_p). The dashed gray line is at 0 on the *x* axis. *P* values smaller than 10^−50^ are capped at that value. **b**, Receiver operating characteristic (ROC) curve (thick black line) for predicting BED in a stratified test set consisting of 10% of the data. The *x* axis shows the false positive rate and the *y* axis shows the true positive rate. The dashed gray line represents chance performance. The area under the curve (AUC) is 0.97. **c**, Precision recall (PR) curve (thick black line) for predicting BED in a stratified test set containing 10% of the data. The *x* axis shows the recall rate and the y axis shows the precision. Positive predictive value (PPV) is 0.11 with a phenotype prevalence of 0.001. The dashed gray line represents change performance.

We tested our model against a holdout group comprising 10% of the cohort and achieved a strong predictive performance: the sensitivity-specificity area under the curve was 97.1% (Fig. 1b), and, while our prevalence of identified BED cases was only 0.1%, the average positive predictive value was 11.0% (Fig. 1c). Thus, we leveraged the extensive information within the MVP EMR to classify undiagnosed but highly probable cases of BED from a smaller cohort of individuals with clinician-diagnosed BED using a hypothesis-free supervised model that relied on many factors previously associated with BED.

## Genetic Architecture of BED independent of BMI

A strong genetic correlation has been reported between a BED-inclusive phenotype and BMI^8^. Therefore, we sought to examine the genetic underpinnings of BED while controlling for BMI by leveraging our machine learning model-derived (MD) BED scores to perform ancestry-specific GWAS on the African ancestry (AFR; *n* = 77,574; Fig. 2a) and European ancestry (EUR; *n* = 285,138; Fig. 2b) populations within the MVP: AFR-MD-BED*BMI, EUR-MD-BED*BMI (Table 1). From our EUR-MD-BED*BMI GWAS, we discovered two genome-wide significant loci at the *HFE* gene (*p* = 1.88×10^−9^) and nearest to the *MCHR2* (*p* = 5.57×10^−10^) gene (Fig. 2b, Table 1). One of the secondary SNPs at the *HFE* locus, rs1800562, corresponds to the C288Y missense mutation with pathogenicity for hemochromatosis. *MCHR2* is a G protein-coupled receptor for melanin-concentrating hormone, a conserved cyclic peptide involved in eating behavior and metabolic homeostasis ^26^. Next, we utilized MAGMA^16^ to identify protein coding genes associated with the EUR-MD-BED*BMI phenotype and found a correlation between the EUR-MD-BED*BMI phenotype and the *APOE* gene (*p* = 7.03×10^−7^). We calculated heritability using LD-Score (LDSC) Regression for the EUR-MD-BED*BMI GWAS to assess the amount of variation explained by an additive SNP model and attained an *h*^2^ of 2.14% (SE = 0.23%, *p* = 6.74×10^−21^). No loci achieved genome-wide significance in the AFR-MD-BED*BMI as it was relatively underpowered (Fig. 2b, Table 1).

**Table 1.** Genome-wide significant loci for MD-BED*BMI GWAS.

| SNP | Chr | Position | Closest Gene | Reference allele | Effect allele | AFR-MD-BED*BMI |  |  | EUR-MD-BED*BMI |  |  | Fixed-MD-BED*BMI |
| --- | --- | --- | --- | --- | --- | --- | --- | --- | --- | --- | --- | --- |
|  |  |  |  |  |  | EAF | Beta (s.e.) | P | EAF | Beta (s.e.) | P | P |
| rs79220007 | 6 | 26098474 | HFE | T | C | 0.011 | 0.023 (0.017) | 0.22 | 0.064 | 0.021 (0.004) | 1.9 x 10<sup>-9</sup> | 8.8 x 10<sup>-10</sup> |
| rs17789218 | 6 | 100600097 | MCHR2 | T | C | 0.045 | 0.007<br>(0.009) | 0.41 | 0.249 | 0.013 (0.002) | $5.6 \times 10^{-10}$ | $4.7 \times 10^{-10}$ |
| rs2275046 | 6 | 150157001 | LRP11 | A | G | 0.804 | 0.016<br>(0.005) | $8.3 \times 10^{-4}$ | 0.358 | 0.009 (0.002) | $1.3 \times 10^{-6}$ | $1.1 \times 10^{-8}$ |

**Abbreviations:** Chr: chromosome; EAF: effect allele frequency; s.e.: standard error

**Fig. 2:**
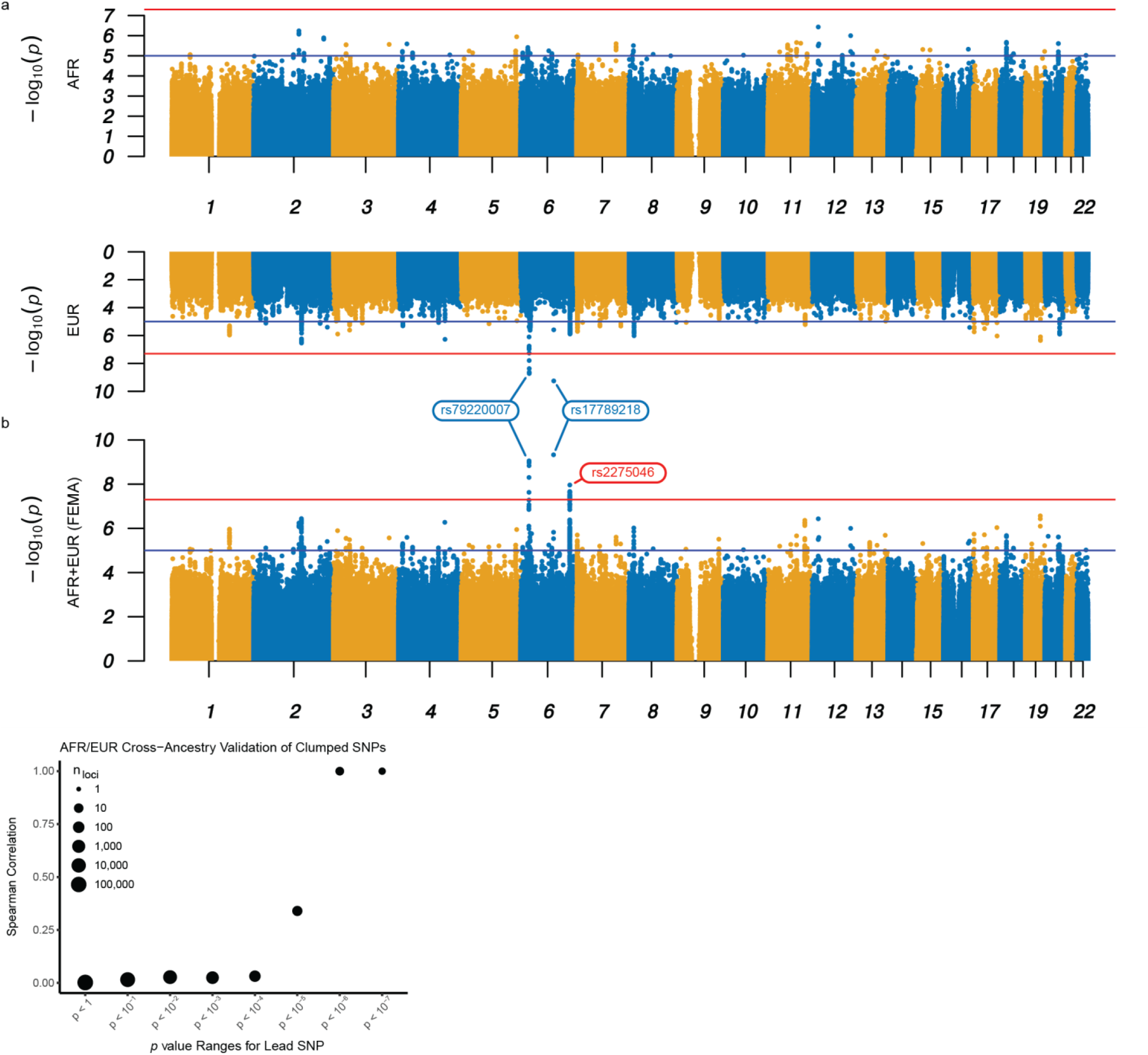
Bi-ancestral GWAS of BED. **a-b**, Miami plot for the AFR-MD-BED*BM (top) and EUR-MD-BED*BMI (bottom) GWAS (a); Manhattan plot for the FEMA-MD-BED*BMI GWAS (b). The *x* axis denotes the chromosome and position of the corresponding SNP. The strength of the SNP-phenotype association is on the *y* axis as the negative log_10_ of uncorrected two-sided *P* value (-log_10_p). The red lines represent genome-wide significance (*p* = 5.0×10^−8^). The blue lines represent the suggestive genome-wide association threshold (*p* = 5.0×10^−5^). Genome-wide significant hits shared by EUR and FEMA GWAS are labeled blue; the unique genome-wide significant hit in FEMA is labeled red. **c**, Correlation between the effect sizes of AFR-MD-BED*BMI and EUR-MD-BED*BMI with progressive restriction of the SNP inclusion threshold. The Spearman correlation for clumped SNPs is shown on the y axis. The threshold below which lead SNPs were included in the correlation analysis is shown on the *x* axis as the uncorrected two-sided *P* value. The size of the point denotes the log_10_ count of the included loci.

We then performed cross-ancestral validation of our genome-wide significant SNPs from the EUR-MD-BED*BMI GWAS. For rs17789218, located near *MCHR2*, we found a consistent effect in the AFR-MD-BED*BMI GWAS (Table 1), but the replication was not significant due to low power. Additionally, we could not replicate the results for the lead SNP at the *HFE* locus, rs79220007, as the minor allele frequency was substantially lower in the AFR (1.1% vs. 6.4% in EUR) cohort and, therefore, the replication was underpowered. Nevertheless, the effect sizes for all SNPs at the *HFE* locus from the AFR cohort had the same sign and similar magnitude to those from the EUR cohort (Table 1). Looking beyond the genome-wide significant loci, we computed Spearman correlation coefficients for clumped SNPs between the EUR and AFR GWAS across the whole genome. We plotted the Spearman correlation between the EUR and the AFR cohorts’ effect sizes for lead SNPs using a series of *p* value thresholds as computed from our EUR-MD-BED*BMI GWAS (Fig. 2d). By limiting our analysis to SNPs with negative log_10_ *p* values below 5, we obtained a correlation of 0.33 between the AFR- and EUR-MD-BED*BMI GWAS. When we further restricted our selection of SNPs to those with lower *p* values, we found progressively increasing correlations from 0.44 to 1 while adjusting the negative log_10_ *p* value threshold from 5.5 to 7.0. As we progressively restricted our analysis to those SNPs with the strongest association to BED in the EUR-MD-BED*BMI GWAS, the increased correlation between the AFR and the EUR MD-BED*BMI GWAS beta coefficients suggest a common genomic signal for BED shared by the EUR and AFR ancestries.

To investigate further the cross-ancestral genetics of BED, we conducted fixed-effects meta-analysis^27^ from the summary statistics of the two ancestry-specific GWAS (FEMA-MD-BED*BMI; Fig. 2; Table 1). In the FEMA-MD-BED*BMI, both of our lead SNPs from the genome-wide significant loci identified in the EUR-MD-BED*BMI GWAS and one additional loci, associated with *LRP11*, achieved genome-wide significance (Table 1). We also performed a Multi-Ancestry Meta-Analysis^28^ and found similar results (MAMA-MD-BED*BMI; Supplementary Fig. 1).

We attempted fine-mapping^29^ to better identify causal/effect variants; however, our findings were limited by the extended linkage disequilibrium blocks within which the index SNPs for the *LRP11* and *HFE* loci reside, the latter falling within the major histocompatibility complex (MHC) locus. Fine-mapping of the locus near *MCHR2* was constrained by the limited coverage quality of this region by the MVP genotyping arrays (Supplementary Table 4).

## GWAS validation

After cross validating our results between the EUR and the AFR cohorts, we performed additional internal and external validations using only the EUR ancestry cohort due to inadequate numbers of AFR ancestry subjects in the validation cohorts. As internal validation, we tested the relationship between our primary GWAS methodology (MD-BED), BMI and an alternative ICD-based BED phenotype (ICD-BED) comprised of individuals reliably diagnosed with BED and/or diagnosed with an ambiguous eating disorder and a with a high probability of having BED based on a second machine learning algorithm designed to discriminate BED from other eating disorders (Methods).

We computed the genetic correlation across five BED-related GWAS on EUR individuals in the MVP: Model-Derived BED (EUR-MD-BED; *n =* 285,138), Model-Derived BED adjusted for BMI (EUR-MD-BED*BMI; *n =* 285,138), ICD-based BED (EUR-ICD-BED; *n =* 549 cases, *n =* 284,648 controls; Supplementary Fig. 2a), ICD-based BED adjusted for BMI (EUR-ICD-BED*BMI; *n =* 549 cases, *n =* 284,648 controls; Supplementary Fig. 2b) and BMI (EUR-BMI; *n =* 291,593). We observed reasonably high correlations between the EUR-ICD-BED GWAS and the other GWAS, with Pearson’s *r* ranging from 0.48 to 0.86 (Fig. 3a). We note that the EUR-MD-BED*BMI GWAS achieved high genetic correlation (*r*_*g*_ = 0.85) with the EUR-ICD-BED*BMI GWAS. In contrast, when we did not adjust our MD-BED phenotype for BMI, we observed that EUR-MD-BED GWAS achieved greater genetic correlation with BMI (*r*_*g*_ = 0.93) than it did with any of the BED GWAS (*r*_*g*_ = 0.71 with EUR-ICD-BED). These results confirm that the two approaches to classification of BED identify common genetic factors, but suggest that without adjustment, our machine learning approach inflated the influence of BMI-associated genetic determinants.

**Fig. 3:**
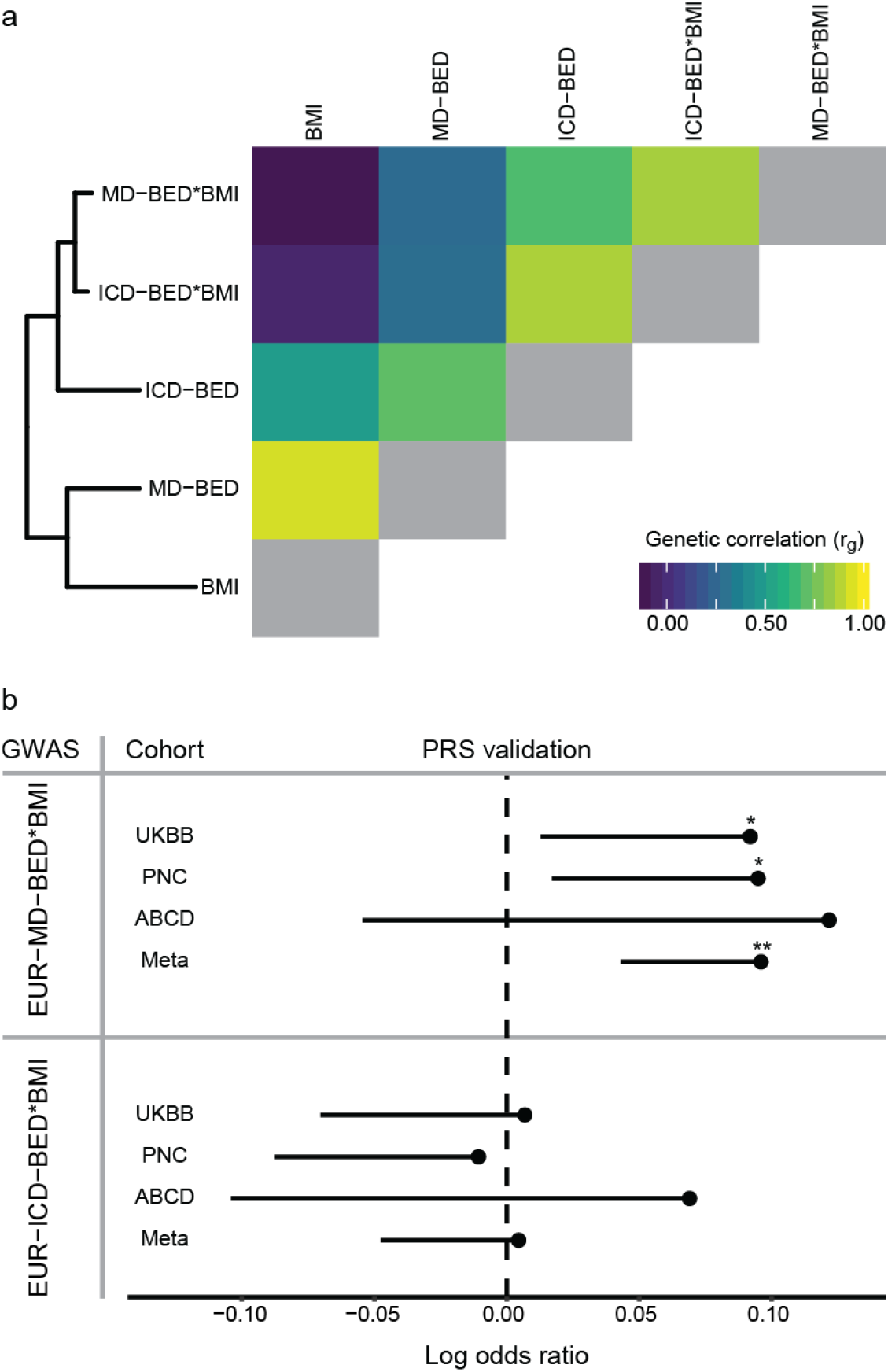
Validation of the MD-BED phenotype. **a**, At the top is a hierarchical clustering of five EUR-BED related phenotypes. On the bottom is a heat map of the genetic correlation matrix. The diagonal genetic correlation entries in yellow represent a correlation of 1 between each GWAS and itself. **b**, PRS validation of EUR-MD-BED*BMI and EUR-ICD-BED*BMI GWAS with UKBB, PNC, ABCD and a meta-analysis of those cohorts. The MVP (vertical) and external (horizontal) cohorts are shown on the *y* axis. The log odds ratio for the PRS predictor is shown on the *x* axis. Confidence intervals are one-sided standard errors. \**p* < 0.05. \*\**p* < 0.01.

While the genetic correlations between the EUR-MD-BED*BMI GWAS and the two ICD-BED GWAS were high, the ICD-based approach identified different genome-wide significant loci (Supplementary Results; Supplementary Table 5). However, heritability of the EUR-ICD-BED GWAS was only nominally significant (*h*^2^ = 22.3%-29.5%, *p* = 0.05) and heritability of the EUR-ICD-BED*BMI GWAS was not significant (*h*^2^ = 16.9%-22.4%, *p* = 0.11).

We also validated our EUR-MD-BED*BMI approach using three external cohorts with BED-inclusive phenotypes by leveraging our GWAS summary statistics to compute PRS. In the UKBB^30^, we defined a BED-inclusive phenotype (*n* = 461; prevalence 0.1%) using the mental health questionnaire and included subjects who self-reported a diagnosis of “psychological over-eating or binge-eating,” while excluding subjects with self-reported diagnoses or inpatient hospital ICD-9/ICD-10 codes of AN or BN. Controls comprised those answering no to these three eating disorder questions and those without ICD codes corresponding to AN or BN (*n* = 385,624; Supplementary Table 6). For the PNC, we identified individuals with a BED-inclusive phenotype based on questions modified from the Kiddie-Schedule for Affective Disorders and Schizophrenia (K-SADS)^18^: we included subjects who reported a history of out-of-control eating and denied compensatory behaviors (e.g. purging; *n* = 531, prevalence 10.9%). Controls comprised those who answered no to the binge-eating and purging questions and screened negative for AN (*n* = 4,330; Supplementary Table 6). For the ABCD, diagnosis of BED was obtained using the K-SADS (*n* = 94, prevalence 2%). Controls comprised those who did not receive a diagnosis of BN (*n* = 4,565; Supplementary Table 6); there was insufficient information to identify subjects with AN. Our EUR-MD-BED*BMI PRS significantly predicted BED in the UKBB (*p* = 0.03) and in the PNC (*p* = 0.02; Fig. 3b), but did not reach significance in the ABCD cohort (*p* = 0.13, Fig. 3b). However, the effect sizes were similar in all validation sets (log odds ratios: 0.09-0.12) and the inverse variance-weighted meta-analysis across the UKBB, ABCD and PNC was robust (*p* = 1.39×10^−3^; Fig. 3). The EUR-ICD-BED*BMI PRS failed to validate in any of the individual cohorts or through meta-analysis (Fig. 3), likely due to the low power and non-significant heritability of this GWAS.

## Shared genetic architecture between BED and other traits

To investigate the genetic overlap between BED and other traits, we computed the genetic correlation between the EUR-MD-BED*BMI GWAS and a set of psychiatric disorders, behavioral phenotypes and health-related traits and contrasted it with the results of our EUR-BMI (Fig. 4a; Supplementary Fig. 3). We found significant positive genetic correlations between EUR-MD-BED*BMI and lobar intracerebral hemorrhage (*r*_*g*_ = 0.56, *p* = 2.02×10^−6^), depression (*r*_*g*_ = 0.52, *p* = 2.46×10^−45^), bipolar disorder (*r*_*g*_ = 0.42, *p* = 7.63×10^−21^), neuroticism (*r*_*g*_ = 0.38, *p* = 5.43×10^−10^), attention-deficit/hyperactivity disorder, (*r*_*g*_ = 0.34, *p* = 1.26×10^−7^), schizophrenia (*r*_*g*_ = 0.24, *p* = 2.94×10^−9^) and AN (*r*_*g*_ = 0.21, *p* = 4.00×10^−4^), as well as a cross-psychiatric disorder GWAS (*r*_*g*_ = 0.44, *p* = 1.58×10^−11^). The strength of the genetic correlation between these traits was greater with the EUR-MD-BED*BMI GWAS than with the EUR-BMI GWAS (Supplementary Fig. 3). We found significant negative genetic correlations between EUR-MD-BED*BMI and educational attainment (*r*_*g*_ = -0.11, *p* = 0.001), intelligence quotient (*r*_*g*_ = -0.12, *p* = 0.019), cognitive performance (*r*_*g*_ = -0.13, *p* = 8.00×10^−4^) and subjective well-being (*r*_*g*_ = -0.21, *p* = 0.006). To validate the negative genetic association between BED and cognitive functioning, we assessed the relationship between our EUR-MD-BED*BMI PRS and neurocognitive measures obtained in the UKBB^31^ and found strong negative associations across nearly all domains tested (Supplementary Fig. 4).

**Fig. 4:**
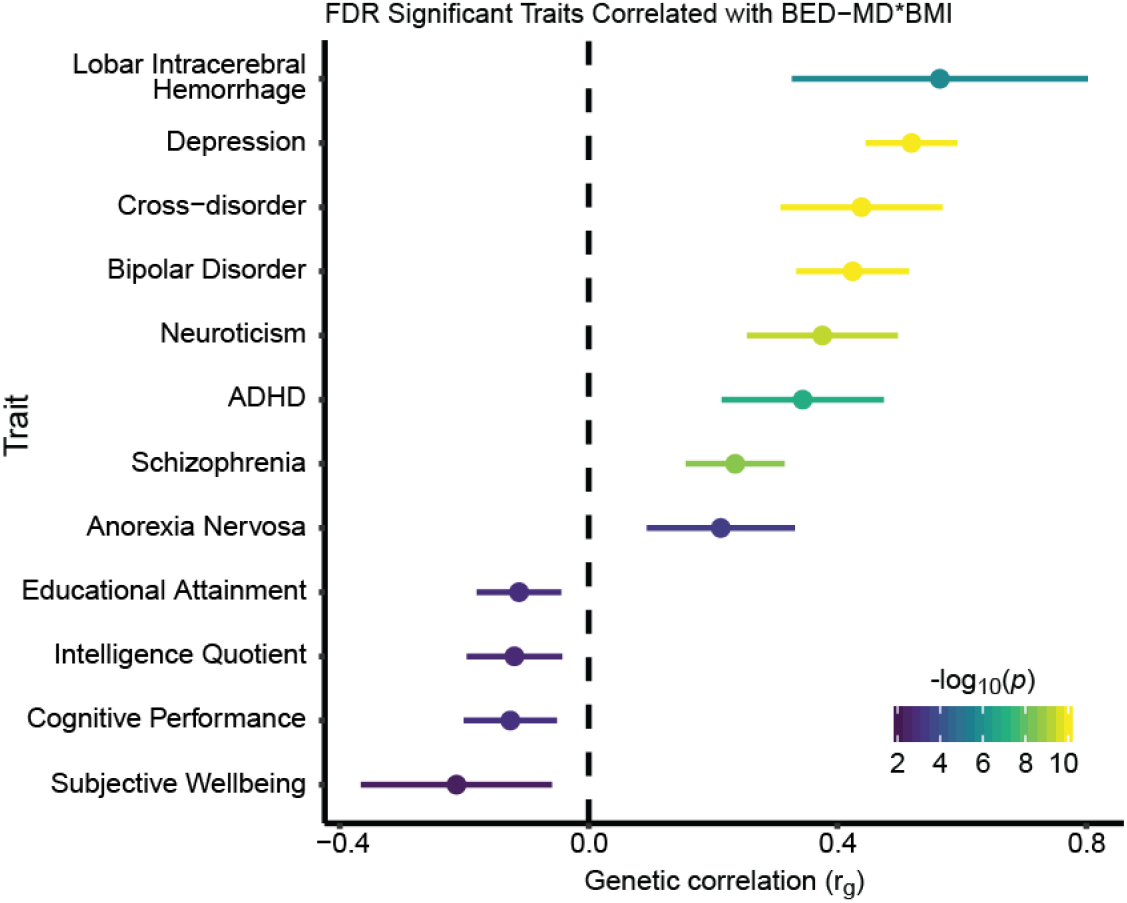
Genetic correlation with other traits. All traits with significant genetic correlation to EUR-MD-BED*BMI at the FDR significant threshold (*q* < 0.05) are ranked by *r*_g_ on the *y* axis. The strength of the genetic correlation is shown on the x axis as r_g_ with the 95% confidence interval for each trait shown and through the color of the error bar corresponding to the uncorrected two-sided *P* value. *P* values smaller than 10^−10^ are capped at that value.

## Pathways and cell types

We used FUMA, excluding the MHC to ensure that results were not driven by the high LD structure and gene density in this region, to identify enrichment in the overlap of gene sets from our EUR-MD-BED*BMI and a large database of published GWAS (Supplementary Fig. 5). Apart from enrichment for neuropsychiatric, obesity-related, autoimmune and cancer traits, we found enrichment for gene sets involved in heme metabolism and biosynthesis and uric acid metabolism. Phenome-wide association studies (PheWAS) of the lead SNPs from the FEMA-MD-BED*BMI GWAS were also consistent with iron dysregulation through associations with disorders of iron metabolism and iron deficiency anemia (Supplementary Fig. 6).

To elucidate the role of iron and heme metabolism in BED, we assessed the relationship between the PRS from our EUR-MD-BED*BMI and EUR-BMI GWAS with both an iron deficiency and an iron overload phenotype derived from the UKBB (Fig. 5a). BED was very strongly positively correlated with iron overload (*β* = 0.66, *p* = 1.62×10^−60^) and negatively correlated with iron deficiency (*β* = -0.025, *p* = 0.01). In contrast, BMI was positively correlated with iron deficiency (*β* = 0.05, *p* = 1.03×10^−7^) and was not significantly correlated with iron overload (*β* = -0.01, *p* = 0.73). To test for a putative causal relationship between BED and iron overload we performed Generalized Summary based-data Mendelian Randomisation (GSMR)^32^, where we leveraged summary statistics from our EUR-MD-BED*BMI and an extant transferrin saturation GWAS^33^ based on an independent cohort comprising subjects from deCODE genetics and the INTERVAL study (Fig. 5b). From the GSMR analysis, we confirmed a statistically robust relationship (*β* = 0.02, *p* = 5.45×10^−6^), further supporting that transferrin saturation, a biomarker of whole body iron stores, affects the model derived BED scores. After linking BED with iron overload, we hypothesized that if iron/heme metabolism is critical for BED we would also see independent downstream enrichment of EUR-MD-BED*BMI risk variants for regions of the genome regulated by heme. Thus, we tested the relationship between enrichment of BED risk variant-homologs and heme-regulated open chromatin regions from a murine erythroid cell □-estradiol stimulation model^34^. After correcting for multiple comparisons, we found significant enrichment of our BED risk variant-homologues in wild-type mice exposed to □-estradiol (induced high heme state; FDR adjusted *p* = 0.03), but not in mutants with reduced heme expression (Fig. 5c), suggesting enrichment of BED risk variants in heme-regulated open chromatin regions.

**Fig. 5:**
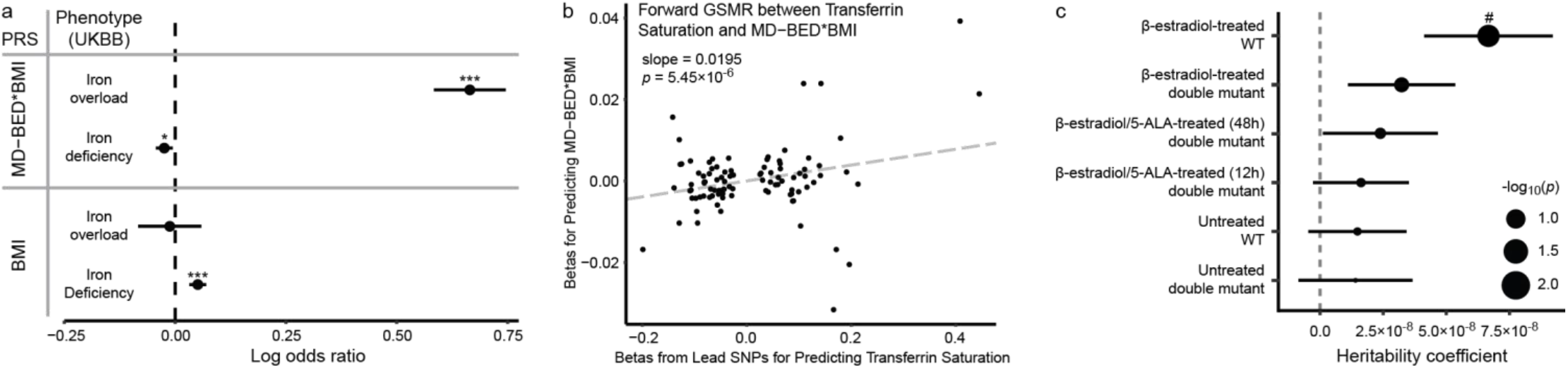
Iron overload in BED. **a**, PRS associations between EUR-MD-BED*BMI and EUR BMI GWAS and iron-related phenotypes in the UKBB. The PRS scores (vertical) and iron phenotypes (horizontal) are shown on the *y* axis. The coefficients, as log odds ratio, from the logistic regression analyses for the PRS predictor are shown on the *x* axis. Confidence intervals are two-sided standard errors. \**p* < 0.05. \*\*\**p* < 0.001. **b**, Scatter plot with generalized linear regression from GSMR between the lead SNPs in the transferrin saturation GWAS from the deCODE genetics and INTERVAL studies and EUR-MD-BED*BMI. Betas from the lead SNPs of the transferrin saturation GWAS are plotted on the *x* axis. Corresponding betas from the EUR-MD-BED*BMI are plotted on the *y* axis. **c**, Enrichment of BED risk variant-homologs in genomic regions of open chromatin in wild type (WT) and heme-deficient mutant murine erythroid cells treated with □-estradiol and/or 5-aminolevulinic acid hydrochloride (5-ALA). Cell lines are represented on the *y* axis. Heritability coefficient is represented on the *x* axis. A positive coefficient signifies enrichment in heritability. Dot size reflects negative log_10_ of uncorrected two-sided *P* value (-log_10_p) of the LD-score regression. Error bars indicate standard errors from the LD-score regression. #*p* < 0.05 after FDR correction.

Based on our gene set enrichment findings, we also attempted to explore the relationship between uric acid metabolism and BED. We found genetic associations with urate and gout; however, in contrast to iron/heme metabolism dysregulation, observed BMI heavily confounded these associations (Supplementary Results; Supplementary table S7).

Finally, to identify cell types in which the genetic drivers of BED are likely to have an effect, we performed partitioned heritability analysis for our EUR-MD-BED*BMI GWAS with two chromatin accessibility atlases^35,36^ and found nominal enrichment across several neural lineages including from limbic system neurons, inhibitory neurons, astrocytes, enteric neurons and enteric glia (Supplementary Results; Supplementary Fig. 7). These results point to a potential pleiotropic effect of the genetic drivers across neural tissues and suggest shared dysfunction across the central and enteric nervous systems.

## Discussion

We report the first GWAS of BED from 362,712 individuals in the MVP, including individuals with AFR and EUR ancestries, and identify three loci that have a genome-wide significant association with BED independent of BMI. We further implicate *APOE* in BED through MAGMA. To power our study, we developed a machine learning approach to identify the probability of individuals having BED across the MVP based on clinician-diagnosed cases. We leveraged the diversity of the MVP to conduct separate GWAS in AFR and EUR cohorts and perform both cross-cohort and meta-analytic validation of our results. Our machine learning approach was effective in performing a GWAS on a common, yet underdiagnosed disorder in a very large cohort. With the limited number of identified cases, our algorithm-based approach shared high genetic correlation (*r*_*g*_ = 0.85) with our more traditional case-control GWAS, while outperforming it on both SNP heritability (*p* = 6.74×10^−21^ vs. *p* = 0.11) and PRS validation across external cohorts (*p* = 1.39×10^−3^ vs. *p* = 0.44).

We found significant association between our MD-BED phenotype built on clinically diagnosed veterans and gender-balanced civilian cohorts. Of the three cohorts, only the ABCD used DSM-based criteria for BED diagnosis; however, it contains only 94 cases as the cohort is substantially younger than the typical age of onset of BED^17^. Thus, the ABCD replication, the only one that did not achieve significant validation, was severely underpowered. The PNC cohort was also younger than the mean age of onset of BED^18^ and the closest available phenotype captured a much larger population than would be expected to have BED. Despite these limitations, our PRS significantly predicted BED in two of the three civilian cohorts (PNC and UKBB) and through meta-analysis. Consequently, we provide strong evidence for the external validity of our GWAS approach, even in the presence of demographic confounders.

A recent UKBB study confirmed the expected partial genetic association between BMI and over-eating/binge-eating in a EUR-only cohort but was not powered to detect associations with individual loci, was not based on diagnosed BED and did not examine the BMI-independent genetic contributions to over-eating/binge-eating^8^. However, their phenotype was similar to the UKBB phenotype that we used as one of the validations of our approach. As expected, we found a high-degree of genetic correlation between BED and BMI using both our machine learning and ICD-based approaches.

While there is substantial epidemiological and genetic overlap between BED and obesity, most obese individuals do not have BED and many people with BED are not obese^3^. Therefore, we examined the genetics of BED while adjusting for BMI. When comparing our EUR-MD-BED*BMI PRS to a number of behavioral and psychiatric traits, we revealed genetic correlations between BED and depression, bipolar disorder and attention-deficit/hyperactivity disorder, all common comorbidities of BED^7^, along with AN. Together, these cross-disorder genetic commonalities provide biological validation of the epidemiological evidence that the BED subpopulation within obesity is a nexus for enhanced psychopathology. We also identified a strong association with intracerebral lobar hemorrhage. This risk may be mediated through shared associated factors including hypertension and through *APOE* mutations^37^.

Examining the individual genes, pathways and cell-types implicated in BED provides further insight into the biology underlying BED and evidence against the theory that BED is merely an associated feature of the co-occurrence of obesity and general psychopathology^38^. Several lines of evidence implicate dysfunctional iron metabolism, heme signaling and resultant iron overload as a driver of BED. We identified multiple SNPs with significant association to BED at the *HFE* gene, including rs1800562, a missense variant pathogenic for hemochromatosis^39^. We identified iron and heme-related phenotypes using both FUMA and PheWAS. We confirmed that BED is genetically associated with iron overload through a PRS association study in the UKBB, transferrin saturation via GSMR in an independent cohort, and further implicated heme metabolism by demonstrating that BED risk variants are enriched for heme-induced open chromatin regions. Heme metabolism has been implicated in insulin resistance^40^ and could partially mediate the association between BED and metabolic dysfunction. Iron deficiency is implicated in pica, another eating disorder in which individuals repetitively eat non-nutritive, nonfood substances^41^. This contrasts with BED in which we implicate iron overload and where patients repetitively engage in consumption of highly nutritive food substances. Iron load may be a partial driver of consumptions on a spectrum from non-nutritive to binge eating through an undiscovered pathway. Iron overload has also been associated with cocaine use disorder^42^, which shares several key features with BED including loss-of-control and impulsivity and animal models have shown an association between binge eating and cocaine craving^43^. Peripheral iron excess may lead to iron accumulation in the basal ganglia, causing oxidative damage and dysfunction of reward circuitry^42^.

We implicate the melanin-concentrating hormone system in BED through the identified genome-wide significant loci near *MCHR2*. Melanin-concentrating hormone plays an important role in energy and glucose metabolism through action at two receptors, MCHR1 and MCHR2^26^, and including regulating impulsive eating ^44^. While MCHR1 is conserved across mammals, MCRH2 exists in only a subset and is less well-studied^26^. Interestingly, a candidate gene study found an association between *MCHR2* and atypical depression, which is characterized by increased appetite/weight gain, hypersomnia and mood reactivity^45^. We also implicate lipid signaling through identification of *LRP11* and *APOE* through GWAS and MAGMA respectively. Validating our genetic approach, obese individuals with BED have an unfavorable lipid profile compared to obese control subjects^46^.

In conclusion, we report the first GWAS in BED and implicate four genes and iron metabolism in the pathophysiology of BED independent of BMI. We demonstrate that BED is a complex, metabolic-psychiatric disorder by inculpating both neural tissues and peripheral metabolic pathways known to influence brain function. Through identification of the melanin-concentrating hormone system and iron metabolism, we find actionable targets for future translational research.

## Online Methods

### Participants

The MVP cohort has been previously described^15,47,48^. Analysis of MVP data was conducted using electronic medical data v20.1 comprising 819,417 patients with demographic information. Of the 819,417 patients, release 3 of the genomics data consists of 459,777 patients with diverse genetic ancestries. Ancestry was assigned using the HARE program^49^ with the 1000 Genomes project^50^ as a reference panel. Confirmatory analysis was performed on the UKBB^31^ (*n* = 386,085), ABCD^17,51^ (*n* = 4,659) and the PNC^18^ (*n* = 4,861), all previously described (Supplementary Table 6).

### Phenotyping

We developed a series of eating disorder phenotype definitions from data available from the subjects’ associated VA electronic medical records available through the MVP. For eating disorder prevalence estimates by diagnostic code within the MVP, we counted subjects that had at least 1 instance of any individual eating disorder coded in either ICD-9-CM or ICD-10-CM (Supplementary Table 1).

We built our BED machine learning model (MD-BED) by leveraging available diagnostic codes, medication prescriptions, BMI measurements and demographic data. The set of reliably diagnosed BED subjects comprised those with either two ICD-10-CM BED codes (F50.81) or a single BED code and no other specific eating disorder codes (Supplementary Table 2). The set of control subjects comprised all MVP subjects with available EMR data, excluding those with any specific eating disorder diagnosis code (Supplementary Table 2) and those who self-reported having an eating disorder on the enrollment questionnaire (“Do you have an eating disorder?”).

We trained a machine learning model to calculate a BED probability score across the MVP utilizing the cohort of reliably diagnosed BED cases described above (*n* = 822). We first mapped ICD diagnoses to Clinical Classification Software codes and computed log counts for diagnoses for each subject. Next, we computed 25th, 50th and 75th percentile BMI measures using height and weight values from each record. We removed records with nonsensical height, weight and BMI values, where we required BMI to range from 9 to 150 kg/m^2^, median height to range from 35 to 95 inches, and weight entries to range from 40 to 1400 pounds. We further removed patients with less than two valid, distinct BMI measurements. Of the 767,527 patients that passed these filters, we constructed a 90%-10% split of our data to train our model. Due to the large number of medication ingredients (3,456) available for analysis, we used a LASSO regression model (using the cv.glmnet function with parameters alpha = 1, family = binomial and nfolds = 10) prior to the creation of the final model to reduce the number of the medication ingredients included. For this step, we took a random sample of 90,000 control subjects to identify the relevant medications, while retaining all of our cases from the training set. To compute the area deprivation index for patients with missing zip codes, we imputed the median value. Subjects without any prescriptions or diagnoses were filtered out. We constructed our final MD-BED model by incorporating log counts of medications at the ingredient level, demographic variables at enrollment (self-reported sex, age, race, and ethnicity, and zipcode-associated deprivation index), log counts for diagnoses and BMI (as calculated above) as model predictors and trained it on the remaining subjects. Ranking of top variables was based on *p* values from the analogous unpenalized logistic regression model, trained on all of the 767,527 patients using only the variables that were retained in the LASSO regression model. The ranking of top variables was only performed to identify important predictors in the model and did not impact downstream analyses.

To construct our MD-BED*BMI model, after assessing the performance of the LASSO logistic regression model for various parameter choices through cross-validation, we initially retained both the model with the lowest binomial deviance (Min) as well as the simplest model that had a mean binomial deviance within one standard deviation of the best performing model (1SE) to avoid overfitting. We then completed GWAS (as described below) on both models adjusting for BMI and retained the model with mean binomial deviance within one standard deviation of the best performing mode (MD-BED*BMI) as it outperformed the model with the lowest binomial deviance (MD-Min-BED*BMI) in SNP heritability (2.14% (s.e. = 0.23%) compared to 1.65% (s.e. = 0.22%)) and did not differ on predictive performance (Supplementary Fig. 8).

For all GWAS utilizing BMI (i.e. EUR-BMI), participant BMI quartile was calculated as above and all participants with distinct and valid BMI measurements at the 25th and 75th percentile were retained.

For privacy reasons, the MVP places a restriction on case-control GWAS requiring at least 500 cases for the summary statistics to be shared with the scientific community for reproducibility, and there are less than 500 individuals of a single ancestry with both an ICD-10-CM BED code and available genetic information. Therefore, to identify enough BED-probable subjects to perform an ICD-based case-control GWAS for the EUR-ICD-BED phenotype, we developed a second machine learning classifier to identify probable BED cases from within the subpopulation of all patients with eating disorder diagnoses. To generate the ICD-BED score, we followed the same procedure as above with cases defined as all reliably diagnosed BED cases. We again built a LASSO regression model, this time training it on the entire cohort of reliably diagnosed BED subjects and training it to distinguish them from within the subpopulation of all subjects with any eating disorder diagnosis. Cases for the GWAS were then then defined as the set of reliably diagnosed subjects with BED as above plus subjects with at least one ambiguous eating disorder code (307.50, 307.59, F50.89, F50.9) and a within-eating disorders BED score greater than the 25 percentile of reliably diagnosed BED cases (*n* = 549). Controls were defined as all remaining subjects excluding those with any eating disorder diagnosis code and those who self-reported having an eating disorder as above (*n* = 284,648).

For the UKBB, inclusion and exclusion was based on the Mental Health Questionnaire and associated ICD-9 and ICD-10 codes. Subjects reporting a “diagnosis of psychological over-eating or binge-eating” were included as cases. Those with a self-reported or ICD coded diagnosis of AN or BN were excluded from both the case and control groups. All remaining subjects were retained as controls. For ABCD (data release 4.0), case-control definitions were based on the K-SADS. Cases were subjects who received a K-SADS diagnosis of BED and controls were subjects who did not receive a K-SADS diagnosis for either BED or BN. A diagnosis of AN could not be made from the available data from ABCD as it could not be determined whether an individual met all criteria or had been formally diagnosed. For the PNC, case-control definitions were based on a modified version of the K-SADS (third study accession, dbGaP phs000607.v3.p2). Cases were those who answered yes to EAT007 (lifetime binge eating) and no to EAT008 (lifetime purging after a binge-eating episode). Subjects were excluded from the control group if they answered yes to EAT007 or EAT008 or screened positive for an AN-like phenotype (yes to EAT001, EAT002 & EAT003 AND either 1) male 2) no to EAT004 3) no to EAT005 or 4) yes to EAT004, EAT005 and EAT006).

In the UKBB, the gout, iron overload, and iron deficiency phenotypes were defined by the presence of at least one relevant phecode or self-reported diagnosis code. Hospital inpatient ICD-9 and ICD-10 codes were converted to phecodes using previously established methods.^52,53^ Gout phenotype cases included individuals with phecodes 274.1 (gout) or 274.11 (gouty arthropathy), as well as self-reported gout at the baseline assessment. Iron overload phenotype cases included individuals with phecodes 275.1 (disorders of iron metabolism) or 277.1 (disorders of porphyrin metabolism). Iron deficiency phenotype cases included individuals with phecodes 262 (mineral deficiency NEC) or 280.1 (iron deficiency anemias, unspecified or not due to blood loss), as well as self-reported iron deficiency anemia at baseline.

### Genotyping, quality control and imputation

Genotyping, quality control and imputation in MVP is handled by a dedicated data team The MVP v3.0 data release used in this study includes genotyping data from 455,789 individuals; DNA was extracted from whole blood (which was collected during enrollment to the MVP) and genotyping was performed with the MVP 1.0 Genotyping array^15^. Ancestry (EUR) as determined by HARE (Harmonized ancestry and race/ethnicity) analysis^49^. Prephasing was performed using EAGLE v2^54^ and genotypes were imputed using Minimac v3 with the 1000 Genomes Project phase 3, version 5 reference panel^55^. Relatedness between MVP participants was inferred using kinship coefficient calculated using software KING^52^. Related individuals are removed using a kinship coefficient cut off >= 0.0884.. PCA to generate ancestral PCs was performed using EIGENSOFT v.6^56,57^.

### Genome-wide association studies

GWAS analysis was conducted employing either logistic regression for binary traits (case-control definition of BED from the ICD model) and linear regression for continuous traits (BMI, MD-BED scores) using PLINK 2.0. We conducted separate analyses for AFR and EUR genetic ancestries and included age, sex, record density and the top 10 principal components to adjust for potential confounders. SNPs with minor allele frequency less than 0.5% or an effective minor allele count < 30 were removed from the analysis per MVP regulation. SNPs with a HWE *p* value less than 5×10^−8^ or an imputation *r*^2^ < 0.4 were removed from the analysis. To assess whether a SNP was genome-wide significant, we used the standard multiple-testing correction threshold, *p* < 5×10^−8^. To satisfy the model assumptions of performing linear regression, we applied an inverse-rank normal transformation of the MD-BED scores to ensure the prediction errors follow an approximately normal distribution for all model derived GWAS.

### Enrichment Analyses

We utilized FUMA (v1.3.7)^58^ and its implementation of MAGMA (v1.0.8)^16^ to determine genes and gene sets that are linked to the MD BED phenotype. We used a Bonferroni-corrected *P* value threshold of 2.684×10^−6^ to account for multiple testing of 18,626 protein coding genes using the 10k UKBB European reference panel (release 2b) to account for linkage disequilibrium. In addition, we considered 54 tissue types from GTEx V8 to assess for gene enrichment across different tissues. We employed the default parameters for excluding annotations from the MHC region, between the *MOG* and *COL11A2* genes.

### LD Score Regression, Heritability and Genetic Correlation

We estimated SNP heritability and genetic correlation using the LDSC package^59,60^. Heritability for case-control phenotypes were computed on the liability scale for a range of plausible population prevalence, while continuous phenotypes were computed on the observed scale. To compute the heritability and genetic correlation estimates, the 1000 Genomes Project -European Project was used to construct the reference panel.

### Meta-Analysis

We used the inverse-variance weighting method to meta-analyze both the AFR and EUR MD-BED*BMI GWAS and the log odds ratio for the PRS on the external cohorts (UKBB, ABCD and PNC). To account for the LD structure and the possibility of different effect sizes between different populations, we also employed Multi-ancestry Meta-analysis (MAMA)^28^ using 1000 Genomes Project as a reference panel. For meta-analysis with MAMA, we filtered out SNPs with minor allele frequencies that were smaller than 1% to ensure our sample size from the reference panel was large enough to attain sufficiently accurate linkage disequilibrium scores. MAMA did not yield any additional hits and as such was not discussed in the main paper.

### Fine-mapping

For each of our loci, where we have a genome-wide significant hit and the minor allele frequency is sufficiently large (> 1%), we performed fine-mapping to identify candidate causal variants using SUSIE with the UKBB as a reference panel. We ran SUSIE^29,61^ both including and excluding outlying SNPs flagged by the algorithm, where we specified the upper bound for the number of causal variants to be 5 and the window to span 1 megabase pairs.

### Phenome-Wide Association Studies

PheWAS were conducted leveraging EUR subjects in the MVP (*n* = 296,407). We then filtered imputed genotypes by minor allele frequency (> 0.01), variant level missingness (< 0.02) and imputation R2 (> 0.9). Phenotypes were derived by aggregating ICD Codes in the EMR data using the categorizations provided in the Phecode Map v1.2^62^; phenotypes with less than 500 cases were removed from the analysis. We then performed a logistic regression, adjusted for sex, age and top 10 ancestry PC’s, to assess the presence of an association between the flagged SNP and the phenotype. We assessed significance using a Bonferroni corrected two-sided *p* value at the 0.05 level.

### Polygenic Risk Scores and Validation

The quality control (QC) and population stratification steps performed on UKBB genotypes used for PRS generation have been described elsewhere^63^. Following these steps, 387,392 samples with European ancestry and 557,369 variants were retained. Processing and imputation performed on the PNC genotypes has also been previously characterized, and resulted in 4,973 European ancestry samples and 4,903,082 variants retained^64^. The ABCD Data Analysis, Informatics & Resource Center (DAIRC) performed initial genotype QC according to the Ricopili pipeline recommendations, resulting in 11,099 samples and 516,598 variants retained in data release 3.0^65^. ABCD genotypes were then lifted-over to GRCh38 using PLINK 1.9 and merged with 1000 Genomes Project genotypes for population stratification^50^. PLINK 2.0 was used to calculate principal components on the merged genotypes, following filtering (minor allele frequency ≥ 0.01, HWE *p* value < 1 × 10^−10^, variant-level missingness ≤ 0.01, regions with high LD removed) and pruning (--indep-pairwise 1000 10 0.02) steps. An ellipsoid with a radius of three standard deviations was calculated from the first three PCs of the 1000 Genomes Project European superpopulation, and ABCD samples that fell outside of this ellipsoid were removed. KING kinship coefficients were calculated in PLINK 2.0 on filtered (minor allele frequency ≥ 0.05, variant-level missingness ≤ 0.1, regions with high LD removed) and pruned (--indep-pairwise 50 5 0.2) genotypes, and only unrelated individuals (KING coefficient ≤ 0.125) were retained^52^. Finally, samples genotyped on plate 461 were removed (as recommended by the ABCD DAIRC team). This resulted in a final sample of 4,676 European ancestry ABCD genotypes.

The Polygenic Risk Score-Continuous Shrinkage (PRS-CS)^66^ method was used to compute PRS for the external cohorts from the GWAS summary statistics derived from the MVP cohort. The European LD reference panel used was generated from UKBB data. We set the parameters as follows: parameter a in the γ-γ prior = 1, parameter b in the γ-γ prior = 0.5, MCMC iterations = 1000, number of burn-in iterations = 500, and thinning of the Markov chain factor = 5. In addition, the global shrinkage parameter phi was derived using Bayesian methodologies. We then employed PLINK 2.0 to compute the individual-level PRS. We assessed the association between scaled (mean = 0, sd = 1) PRS and the BED-inclusive phenotype in each cohort through logistic regression, where we adjusted for age, age squared, genotyping batch, sex and ancestry (first 20 principal components obtained from European ancestry samples in UKBB/ABCD, first 10 MDS dimensions obtained from European ancestry samples in PNC). *P* values for PRS were computed using a one-sided hypothesis test. In the UKBB we additionally assessed PRS association with gout, iron overload, and iron deficiency phenotypes using logistic regression, as well as blood urate levels at baseline and neurocognitive phenotypes using linear regression. Urate levels were square-root-transformed, while neurocognitive phenotypes were scaled (mean = 0, sd = 1). Models were generated both with and without BMI as a covariate, and *P* values were computed using a two-sided hypothesis test.

The neurocognitive phenotypes assessed included numeric memory (UKBB data-field 4282), reaction time (data-field 20023, log-transformed and reverse-coded so higher scores indicate better performance), pairs matching (data-field 399, trial 2 only, log+1-transformed and reverse-coded), matrix pattern completion (data-field 6373 divided by data-field 6374), tower rearranging (data-field 21004), numeric trail making (data-field 6348, log-transformed and reverse-coded), alphanumeric trail making (data-field 6350, log-transformed and reverse-coded), symbol digit substitution (data-field 23324), and fluid intelligence/reasoning (data-field 20016). Neurocognitive results recorded at the participants’ first imaging follow-up assessments were used, except in the case of numeric memory, reaction time, pairs-matching, and fluid intelligence/reasoning phenotypes where baseline data was instead used. We adjusted reported *p* values using the false discovery rate to account for the number of tests performed.

### Concordance of EUR and AFR GWAS hits

To assess the similarity between the AFR- and EUR BED-MD*BMI GWAS, we clumped lead SNPs in Plink^67^ using the European 1k Genomes reference panel. In particular, we required that all SNPs in the same clump must have an R^2^ of at least 0.2 with the lead SNP and be at most 250 kbp away from the lead SNP. We then computed the Spearman correlation between the linear regression coefficients for the respective GWAS using different *P* value filters, (e.g. *p* < 10^−4^) as described in the main text.

### Generalised Summary-data-based Mendelian Randomisation

We leveraged the GCTA software tool^68^ to perform GSMR using the European participants from the 1000 Genomes Project as a reference panel with the default parameters. Since GSMR requires at least 10 genome wide significant loci, we could only conduct forward GSMR to assess if a given phenotype (transferrin saturation, baseline urate levels) influences our model derived BED phenotype, and not backward GSMR as our MD-BED GWAS lacks the requisite number of genome-wide significant loci. We used available summary statistics for the transferrin saturation^33^ and urate level^69^ phenotypes. As part of GSMR, pleiotropic SNPs were filtered out using the HEIDI-outlier method before assessing the correlation of lead SNPs from the summary statistics for transferrin saturation phenotype with the model derived BED phenotype.

### Partitioned Heritability

We examined an overlap of common genetic variants of BED and open chromatin from (i) wild-type and heme-deficient mutant murine erythroid cells treated with □-estradiol and/or 5-aminolevulinic acid hydrochloride (5-ALA)^34^, (ii) a sciATAC-seq3 study of human fetal cell types^35^, (iii) a scATAC-seq study across six human adult brain regions^36^ using an LD-score partitioned heritability approach^70^. We used LD-scores with a baseline model of general genomic annotation (such as conserved regions and coding regions) that corrects for the general genetic context of tested sets of open chromatin regions. All regions of open chromatin were extended by 500 base pairs in either direction. The broad MHC-region (chr6:25-35MB) was excluded due to its extensive and complex LD structure, but otherwise default parameters were used for the algorithm. In the case of mouse cell lines, we merged all peaks of open chromatin for all replicates belonging to the same type of experiment (i.e. untreated / □-estradiol treated wild-type, untreated / □-estradiol mutant and untreated / □-estradiol treated double mutant) and converted mice genome coordinates of resulting peak sets to human genome coordinates using liftOver (https://genome.ucsc.edu/cgi-bin/hgLiftOver).

## Supporting information

Strobe Checklist

## Data Availability

Data produced in the present study will be shared through dbGaP upon publication of the final version of the manuscript.

## Data Availability

Data will become available on dbGaP when paper is accepted.

## Online Acknowledgements

This research is based on data from the Million Veteran Program, Office of Research and Development, Veterans Health Administration, and was supported by award #MVP006. This publication does not represent the views of the Department of Veteran Affairs or the United States Government. This study was also supported by the National Institutes of Health (NIH), Bethesda, MD under award numbers T32MH087004 (Karen Therrien), T32MH122394 (Amirhossein Modabbernia), K08MH122911 (Georgios Voloudakis), R01MH125246, R01AG067025, U01MH116442, R01MH109677 (Panos Roussos), and by the Veterans Affairs Merit grants BX002395 and BX004189 (Panos Roussos). This study has also been funded in part by the Brain & Behavior Research Foundation via the 2020 NARSAD Young Investigator Grant #29350 (Georgios Voloudakis). We thank Shing Wan Choi and Paul F. O’Reilly for their guidance and expertise in utilizing data from the UK Biobank. We thank the participants in the UK Biobank and the scientists involved in the construction of this resource. This research has been conducted using the UK Biobank Resource under application 18177 (P.F.O.). This work was supported, in part, through the computational resources and staff expertise provided by Scientific Computing at the Icahn School of Medicine at Mount Sinai. Data used in the preparation of this article were obtained from the Adolescent Brain Cognitive Development (ABCD) Study (https://abcdstudy.org), held in the NIMH Data Archive (NDA). This is a multisite, longitudinal study designed to recruit more than 10,000 children aged 9-10 and follow them over 10 years into early adulthood. The ABCD Study is supported by the National Institutes of Health and additional federal partners under award numbers U01DA041022, U01DA041028, U01DA041048, U01DA041089, U01DA041106, U01DA041117, U01DA041120, U01DA041134, U01DA041148, U01DA041156, U01DA041174, U24DA041123, U24DA041147, U01DA041093, and U01DA041025. A full list of supporters is available at https://abcdstudy.org/federal-partners.html. A listing of participating sites and a complete listing of the study investigators can be found at https://abcdstudy.org/scientists/workgroups/. ABCD consortium investigators designed and implemented the study and/or provided data but did not necessarily participate in analysis or writing of this report. This manuscript reflects the views of the authors and may not reflect the opinions or views of the NIH or ABCD consortium investigators. The ABCD data repository grows and changes over time. Support for data collection for the PNC, acquired through dbGaP (accession no. phs000607, v3.p2), was provided by grant RC2MH089983 awarded to R. Gur and RC2MH089924 was awarded to H. Hakonarson. Participants were recruited and genotyped through the Center for Applied Genomics (CAG) at The Children’s Hospital in Philadelphia (CHOP). Phenotypic data collection occurred at the CAG/CHOP and at the Brain Behavior Laboratory, University of Pennsylvania.

## Notes

### Competing Interest Statement

Drs. Hildebrandt is a scientific advisory board member of Noom, Inc. and Drs. Hildebrandt and Sysko receive funding from and have equity in Noom, Inc (non-publicly traded company). Dr. Sysko receives royalties from Wolters Kluwer Health.

### Author Declarations

The present study, which involved secondary analysis of existing data collected through the primary Million Veteran Program study, was reviewed and approved by the Department of Veterans Affairs Central Institutional Review Board.

