## Supplemental Tables for "Genome-wide analysis of binge-eating disorder identifies the first three risk loci and implicates iron metabolism"

**Table S1 Demographics of eating disorder diagnoses in the MVP**

| Diagnosis | ICD-9CM Codes | ICD-10CM Codes | n | Female | Black or African American | White | Mean Age (sd), years | Mean of Median BMI kg/m^2 (sd) |
|---|---|---|---|---|---|---|---|---|
| Anorexia Nervosa | 307.1 | F50.00, F50.01, F50.02 | 494 | 61.1% | 17.0% | 73.3% | 50.4 (14.8) | 24.8 (6.7) |
| Bulimia Nervosa | 307.51 | F50.2 | 876 | 70.2% | 15.5% | 73.5% | 45.4 (12.8) | 30.2 (7.3) |
| Binge-Eating Disorder | | F50.81 | 851 | 40.5% | 14.4% | 78.8% | 49.3 (12.7) | 38.2 (8.3) |
| Pica | 307.52 | | 55 | 58.2% | 52.7% | 36.4% | 51.4 (11.4) | 30.6 (6.4) |
| Rumination Disorder | 307.53 | | 18 | 11.1% | 11.1% | 88.9% | 62.9 (10.4) | 29.4 (5.4) |
| Avoidant/Restrictive Food Intake Disorder | | F50.82 | 22 | 72.7% | 18.2% | 63.6% | 47.8 (12.7) | 24.1 (5.2) |
| Psychogenic Vomiting | 307.54 | | 14 | 42.9% | 42.9% | 57.1% | 49.6 (12.9) | 32.7 (5.0) |
| Ambiguous Diagnoses | 307.50, 307.59 | F50.89, F50.9 | 2896 | 45.1% | 16.5% | 77.2% | 52.6 (13.8) | 34.1 (9.4) |

**Note:** Demographics are self-report survey data at study entry. BMI is the group mean of subject's median BMI derived from the electronic medical record. Diagnoses are reported where there is at least one ICD-9CM or ICD-10CM diagnosis in the electronic medical record. **Abbreviations:** BMI: body mass index; MVP: Million Veteran Program; s.e.: standard error

**Table S2 Demographics of reliably diagnosed BED cases and controls for BED-score calculation (MD-BED)**

| | BED | Control |
|---|---|---|
| **n** | 822 | 766,705 |
| **Female** | 40.2% | 9.3% |
| **Black or African American** | 14.2% | 19.1% |
| **White** | 79.3% | 74.3% |
| **Mean Age (sd), years** | 49.5 (12.7) | 61.3 (14.3) |
| **Mean of Median BMI (sd), kg/m^2** | 38.3 (8.3) | 30.1 (5.7) |
| **Comorbidities** | | |
| **Anorexia Nervosa** | 2.1% | 0.0% |
| **Bulimia Nervosa** | 5.4% | 0.0% |

**Note:** Demographics are self-report survey data study entry. BMI is the group mean of subject's median BMI derived from the electronic medical record. Combormitidies are reported where there is at least one ICD-9CM or ICD-10CM diagnosis.
**Abbreviations:** BED: binge-eating disorder; BMI: body mass index

| Table S3 Full list of BED prectictors | | | |
|---|---|---|---|
| **Term** | **Estimate** | **CCS Code** | **Variable Type** |
| Sulfanilamide | 4.56 | | Medication ingredient |
| Vidarabine | 3.42 | | Medication ingredient |
| Constriction | 2.02 | | Medication ingredient |
| Antitoxins | 1.92 | | Medication ingredient |
| Foamcoat | 1.88 | | Medication ingredient |
| Pro | 1.74 | | Medication ingredient |
| Penciclovir | 1.33 | | Medication ingredient |
| Estrogren | 1.33 | | Medication ingredient |
| Crofelemer | 1.14 | | Medication ingredient |
| Lisdexamfetamine | 1.05 | | Medication ingredient |
| Miscellaneous mental health disorders | 1.00 | 670 | Diagnosis |
| Female (Gender) | 0.83 | | Demographic |
| Follicle Stimulanting Hormone | 0.76 | | Medication ingredient |
| Lorcaserin | 0.72 | | Medication ingredient |
| Cold Cream | 0.61 | | Medication ingredient |
| Ospemifene | 0.61 | | Medication ingredient |
| Apalutamide | 0.53 | | Medication ingredient |
| Phentermine | 0.44 | | Medication ingredient |
| Butoconazole | 0.41 | | Medication ingredient |
| Elagolix | 0.39 | | Medication ingredient |
| Magnesium Chloride | 0.29 | | Medication ingredient |
| Other nutritional, endocrine and metabolic disorders | 0.28 | 58 | Diagnosis |
| Delirium, dementia and amnestic and other cognitive disorders | -0.15 | 653 | Diagnosis |
| Glucose Sensor | 0.14 | | Medication ingredient |
| Mood disorders | 0.14 | 657 | Diagnosis |
| Non-traumatic joint disorders | -0.13 | 13.2 | Diagnosis |
| Median BMI | 0.11 | | Measurements |
| Black or African American (Race) | -0.10 | | Demographic |
| Liraglutide | 0.09 | | Medication ingredient |
| Dulaglutide | 0.09 | | Medication ingredient |

| | | | |
|---|---|---|---|
| Quetiapine | -0.08 | | Medication ingredient |
| Hypertension | -0.07 | 7.1 | Diagnosis |
| Vortioxetine | 0.06 | | Medication ingredient |
| Lamotrigine | 0.05 | | Medication ingredient |
| Bupropion | 0.05 | | Medication ingredient |
| Anxiety disorders | 0.05 | 651 | Diagnosis |
| Supply | -0.04 | | Medication ingredient |
| Paralysis | -0.04 | 82 | Diagnosis |
| Epilepsy and convulsions | -0.04 | 83 | Diagnosis |
| Disorders of teeth and jaw | -0.04 | 136 | Diagnosis |
| Escitalopram | 0.03 | | Medication ingredient |
| Age | -0.03 | | Demographic |
| Fluoxetine | 0.03 | | Medication ingredient |
| Attention deficit, conduct and disruptive behavior disorders | 0.02 | 652 | Diagnosis |
| Eye disorders | -0.02 | 6.7 | Diagnosis |
| White (Race) | 0.01 | | Demographic |
| Risperidone | -0.01 | | Medication ingredient |
| Intracranial injury | -0.01 | 233 | Diagnosis |
| Naltrexone | 0.01 | | Medication ingredient |
| Trazodone | -0.01 | | Medication ingredient |
| Medication organizer | -0.01 | | Medication ingredient |
| Upper gastrointestinal disorders | -0.01 | 9.4 | Diagnosis |
| Atomoxetine | 0.01 | | Medication ingredient |
| Schizophrenia and other psychotic disorders | -4.38E-03 | 659 | Diagnosis |
| Superficial injury; contusion | -2.29E-03 | 239 | Diagnosis |
| Antivenins | 6.59E-05 | | Medication ingredient |
| Luteinizing Hormone | 5.71E-05 | | Medication ingredient |
| **Abbreviations:** BED: binge-eating disorder; CCS: Clinical Classifications Software; Pro: Progens | | | |

| Table S4 Fine-mapping of lead SNPs | | | | |
|---|---|---|---|---|
| **Lead SNP** | **Chr** | **Position** | **Lead SNP in 95% Credible Set** | **SNPs with Probability of being Causal > 95%** |
| rs79220007 | 6 | 26098474 | No | rs1572982, rs198856, rs198843, rs11465177 |
| rs17789218 | 6 | 100600097 | Yes | rs17789218, rs4839776 |
| rs9322224 | 6 | 150153901 | No | None |

**Note:** Lead SNP is the lead SNP in the EUR-MD-BED*BMI GWAS from each locus identified as a genome-wide significant hit in either the EUR-MD-BED*BMI or Fixed-MD-BED*BMI GWAS **Abbreviations:** Chr: chromosome; GWAS: genome-wide association studies; SNP: single nucleotide polymorphism

**Table S5 Genome-wide significant loci for EUR-ICD-BED and EUR-ICD-BED*BMI GWAS**

| | | | | | | | EUR-ICD-BED | | EUR-ICD-BED*BMI | |
|---|---|---|---|---|---|---|---|---|---|---|
| **SNP** | **Chr** | **Position** | **Prioritized Gene** | **Reference allele** | **Effect allele** | **EAF** | **Beta (s.e.)** | **P** | **Beta (s.e.)** | **P** |
| rs150714684 | 17 | 31648819 | ASIC2 | A | G | 0.016 | 1.147 (0.202) | 1.27x10^(-8) | 1.272 (0.205) | 5.94x10^(-10) |
| rs149704691 | 7 | 8694971 | NXPH1 | C | T | 0.005 | 1.725 (0.335) | 2.64x10^(-7) | 1.902 (0.341) | 2.51x10^(-8) |

**Abbreviations:** Chr: Chromosome; EAF: Effect allele frequency; GWAS: genome-wide association studies; s.e.: standard error; SNP: single nucleotide polymorphism

**Table S6 Demographics of total cohort, included cases and controls for UK Biobank, PNC and ABCD Validation**

| | UKBB | | | PNC | | | ABCD | | |
|---|---|---|---|---|---|---|---|---|---|
| | Full Cohort | BED | Control | Full Cohort | BED | Control | Full Cohort | BED | Control |
| **n** | 502488 | 461 | 385624 | 9496 | 531 | 4330 | 11876 | 94 | 4565 |
| **Female (%)** | 54.4 | 85.2 | 53.8 | 51.5 | 48.2 | 48.5 | 47.8 | 44.7 | 47.1 |
| **White (%)** | 94.0 | 98.5 | 99.2 | 54.6 | 91.9 | 92.7 | 52.0 | 90.4 | 96.8 |
| **Black (%)** | 1.6 | 0.0 | 0.0 | 33.2 | 0.2 | 0.4 | 15.0 | 0.0 | 0.0 |
| **Mean Age (sd), years** | 56.5 (8.1) | 53.3 (7.5) | 56.7 (8.0) | 13.7 (3.7) | 14.3 (3.1) | 13.7 (3.7) | 9.9 (0.6) | 9.9 (0.6) | 9.9 (0.6) |
| **Median BMI kg/m^2** | 26.7 | 32.0 | 26.7 | 21.1 | 23.4 | 20.0 | 17.6 | 22.6 | 17.1 |
| **Mean BMI kg/m^2 (sd)** | 27.4 (4.8) | 32.7 (7.8) | 27.4 (4.8) | 22.4 (6.3) | 24.2 (6.1) | 20.8 (5.0) | 18.7 (4.0) | 22.5 (4.4) | 17.9 (3.2) |
| **Comorbidities** | | | | | | | | | |
| **Anorexia Nervosa (%)** | 0.2 | 0.0 | 0.0 | 1.2 | 0.0 | 0.0 | Not Available | Not Available | Not Available |
| **Bulimia Nervosa (%)** | 0.1 | 0.0 | 0.0 | 0.8 | 0.0 | 0.0 | 0.2 | 0.0 | 0.0 |

**Note:** Demographics are self-report survey data from baseline assessment. BMI is calculated from height and weight measured during the baseline assessment. For UKBB, diagnoses are reported where there is at least one ICD-9 or ICD-10 diagnosis in the electronic medical record, and/or reported in a follow-up mental health survey administered to a subset of participants in 2016. For PNC, diagnoses are reported based on participant and/or parent informant responses to eating disorder-related questions modified from the K-SADS. For ABCD, diagnoses are reported based on participant and/or parent informant responses to the K-SADS. **Abbreviations:** BMI: body mass index; UKBB: UK Biobank; PNC: Philadelphia Neurodevelopmental Cohort; ABCD: Adolescent Brain Cognitive Development; SD: standard deviation; K-SADS: Kiddie-Schedule for Affective Disorders and Schizophrenia

**Table S7 PRS association analysis with uric acid phenotypes in UKBB**

| Phenotype | Phenotype for Generating PRS | Additional Confounder Adjustment | Beta | P |
|---|---|---|---|---|
| Gout | EUR-BMI | No | 0.09 | 7.17*10^(-14) |
| Gout | EUR-BMI | BMI | -0.057 | 4.3*10^(-6) |
| Gout | EUR-MD-BED*BMI | No | -0.037 | 1.72*10^(-3) |
| Gout | EUR-MD-BED*BMI | BMI | -0.034 | 4.88*10^(-3) |
| Urate Levels | EUR-BMI | No | 0.098 | 1.5*10^(-205) |
| Urate Levels | EUR-BMI | BMI | -0.063 | 3.73*10^(-96) |
| Urate Levels | EUR-MD-BED*BMI | No | -0.025 | 1.21*10^(-15) |
| Urate Levels | EUR-MD-BED*BMI | BMI | -0.022 | 1.22*10^(-13) |

**Abbreviations:** BED: binge-eating disorder, BMI: body mass index, EUR: European, MD: model-derived, UKBB: UK Biobank, PRS: polygenic risk score
