## Supplemental Text and Figures for "Genome-wide analysis of binge-eating disorder identifies the first three risk loci and implicates iron metabolism"

### Supplementary Appendix

VA Million Veteran Program Core Acknowledgement **1**

Supplementary Figures **7**

Supplementary Results **18**

**VA Million Veteran Program Core Acknowledgement**

MVP Program Office

- Program Director - Sumitra Muralidhar, Ph.D.
US Department of Veterans Affairs, 810 Vermont Avenue NW, Washington, DC 20420
- Associate Director, Scientific Programs - Jennifer Moser, Ph.D.
US Department of Veterans Affairs, 810 Vermont Avenue NW, Washington, DC 20420
- Associate Director, Cohort Management & Public Relations - Jennifer E. Deen, B.S.
US Department of Veterans Affairs, 810 Vermont Avenue NW, Washington, DC 20420

MVP Executive Committee

- Co-Chair: J. Michael Gaziano, M.D., M.P.H.
VA Boston Healthcare System, 150 S. Huntington Avenue, Boston, MA 02130
- Co-Chair: Sumitra Muralidhar, Ph.D.
US Department of Veterans Affairs, 810 Vermont Avenue NW, Washington, DC 20420
- Jean Beckham, Ph.D.
Durham VA Medical Center, 508 Fulton Street, Durham, NC 27705
- Kyong-Mi Chang, M.D.
Philadelphia VA Medical Center, 3900 Woodland Avenue, Philadelphia, PA 19104
- Philip S. Tsao, Ph.D.
VA Palo Alto Health Care System, 3801 Miranda Avenue, Palo Alto, CA 94304
- Shiuh-Wen Luoh, M.D., Ph.D.
VA Portland Health Care System, 3710 SW US Veterans Hospital Rd, Portland, OR 97239
US Department of Veterans Affairs, 810 Vermont Avenue NW, Washington, DC 20420
- Juan P. Casas, M.D., Ph.D., Ex-Officio
VA Boston Healthcare System, 150 S. Huntington Avenue, Boston, MA 02130

MVP Principal Investigators

- J. Michael Gaziano, M.D., M.P.H.
VA Boston Healthcare System, 150 S. Huntington Avenue, Boston, MA 02130
- Philip S. Tsao, Ph.D.
VA Palo Alto Health Care System, 3801 Miranda Avenue, Palo Alto, CA 94304

MVP Operations

- MVP Executive Director – Juan P. Casas, M.D., Ph.D.
VA Boston Healthcare System, 150 S. Huntington Avenue, Boston, MA 02130
- Director of Regulatory Affairs – Lori Churby, B.S.
VA Palo Alto Health Care System, 3801 Miranda Avenue, Palo Alto, CA 94304
- MVP Cohort Management Director – Stacey B. Whitbourne, Ph.D.
VA Boston Healthcare System, 150 S. Huntington Avenue, Boston, MA 02130
- MVP Recruitment/Enrollment Director - Jessica V. Brewer, M.P.H.
VA Boston Healthcare System, 150 S. Huntington Avenue, Boston, MA 02130
- Director, VA Central Biorepository, Boston – Mary T. Brophy M.D., M.P.H.
VA Boston Healthcare System, 150 S. Huntington Avenue, Boston, MA 02130
- Executive Director for MVP Biorepositories - Luis E. Selva, Ph.D.
VA Boston Healthcare System, 150 S. Huntington Avenue, Boston, MA 02130
- MVP Informatics, Boston – Shahpoor (Alex) Shayan, M.S.
VA Boston Healthcare System, 150 S. Huntington Avenue, Boston, MA 02130
- Director, MVP Data Operations/Analytics, Boston – Kelly Cho, M.P.H., Ph.D.
VA Boston Healthcare System, 150 S. Huntington Avenue, Boston, MA 02130
- Director, Center for Computational and Data Science (C-DACS) & Genomics Core – Saiju Pyarajan Ph.D.
VA Boston Healthcare System, 150 S. Huntington Avenue, Boston, MA 02130
- Director, Molecular Data Core – Philip S. Tsao, Ph.D.
VA Palo Alto Health Care System, 3801 Miranda Avenue, Palo Alto, CA 94304
- Director, Phenomics Data Core – Kelly Cho, M.P.H, Ph.D.
VA Boston Healthcare System, 150 S. Huntington Avenue, Boston, MA 02130
- Director, VA Informatics and Computing Infrastructure (VINCI) – Scott L. DuVall, Ph.D.
VA Salt Lake City Health Care System, 500 Foothill Drive, Salt Lake City, UT 84148
- MVP Coordinating Centers
    - Cooperative Studies Program Clinical Research Pharmacy Coordinating Center, Albuquerque – Todd Connor, Pharm.D.; Dean P. Argyres, B.S., M.S.
    New Mexico VA Health Care System, 1501 San Pedro Drive SE, Albuquerque, NM 87108
    - Genomics Coordinating Center, Palo Alto – Philip S. Tsao, Ph.D.

VA Palo Alto Health Care System, 3801 Miranda Avenue, Palo Alto, CA 94304

o MVP Boston Coordinating Center, Boston - J. Michael Gaziano, M.D., M.P.H.

VA Boston Healthcare System, 150 S. Huntington Avenue, Boston, MA 02130

o MVP Information Center, Canandaigua – Brady Stephens, M.S.

Canandaigua VA Medical Center, 400 Fort Hill Avenue, Canandaigua, NY 14424

Current MVP Local Site Investigators

- Atlanta VA Medical Center (Peter Wilson, M.D.)
1670 Clairmont Road, Decatur, GA 30033
- Bay Pines VA Healthcare System (Rachel McArdle, Ph.D.)
10,000 Bay Pines Blvd Bay Pines, FL 33744
- Birmingham VA Medical Center (Louis Dellitalia, M.D.)
700 S. 19th Street, Birmingham AL 35233
- Central Western Massachusetts Healthcare System (Kristin Mattocks, Ph.D., M.P.H.)
421 North Main Street, Leeds, MA 01053
- Cincinnati VA Medical Center (John Harley, M.D., Ph.D.)
3200 Vine Street, Cincinnati, OH 45220
- Clement J. Zablocki VA Medical Center (Jeffrey Whittle, M.D., M.P.H.)
5000 West National Avenue, Milwaukee, WI 53295
- VA Northeast Ohio Healthcare System (Frank Jacono, M.D.)
10701 East Boulevard, Cleveland, OH 44106
- Durham VA Medical Center (Jean Beckham, Ph.D.)
508 Fulton Street, Durham, NC 27705
- Edith Nourse Rogers Memorial Veterans Hospital (John Wells., Ph.D.)
200 Springs Road, Bedford, MA 01730
- Edward Hines, Jr. VA Medical Center (Salvador Gutierrez, M.D.)
5000 South 5th Avenue, Hines, IL 60141
- Veterans Health Care System of the Ozarks (Kathrina Alexander, M.D.)
1100 North College Avenue, Fayetteville, AR 72703
- Fargo VA Health Care System (Kimberly Hammer, Ph.D.)
2101 N. Elm, Fargo, ND 58102
- VA Health Care Upstate New York (James Norton, Ph.D.)
113 Holland Avenue, Albany, NY 12208
- New Mexico VA Health Care System (Gerardo Villareal, M.D.)
1501 San Pedro Drive, S.E. Albuquerque, NM 87108
- VA Boston Healthcare System (Scott Kinlay, M.B.B.S., Ph.D.)

150 S. Huntington Avenue, Boston, MA 02130

- VA Western New York Healthcare System (Junzhe Xu, M.D.)

3495 Bailey Avenue, Buffalo, NY 14215-1199

- Ralph H. Johnson VA Medical Center (Mark Hamner, M.D.)

109 Bee Street, Mental Health Research, Charleston, SC 29401

- Columbia VA Health Care System (Roy Mathew, M.D.)

6439 Garners Ferry Road, Columbia, SC 29209

- VA North Texas Health Care System (Sujata Bhushan, M.D.)

4500 S. Lancaster Road, Dallas, TX 75216

- Hampton VA Medical Center (Pran Iruvanti, D.O., Ph.D.)

100 Emancipation Drive, Hampton, VA 23667

- Richmond VA Medical Center (Michael Godschalk, M.D.)

1201 Broad Rock Blvd., Richmond, VA 23249

- Iowa City VA Health Care System (Zuhair Ballas, M.D.)

601 Highway 6 West, Iowa City, IA 52246-2208

- Eastern Oklahoma VA Health Care System (River Smith, Ph.D.)

1011 Honor Heights Drive, Muskogee, OK 74401

- James A. Haley Veterans' Hospital (Stephen Mastorides, M.D.)

13000 Bruce B. Downs Blvd, Tampa, FL 33612

- James H. Quillen VA Medical Center (Jonathan Moorman, M.D., Ph.D.)

Corner of Lamont & Veterans Way, Mountain Home, TN 37684

- John D. Dingell VA Medical Center (Saib Gappy, M.D.)

4646 John R Street, Detroit, MI 48201

- Louisville VA Medical Center (Jon Klein, M.D., Ph.D.)

800 Zorn Avenue, Louisville, KY 40206

- Manchester VA Medical Center (Nora Ratcliffe, M.D.)

718 Smyth Road, Manchester, NH 03104

- Miami VA Health Care System (Ana Palacio, M.D., M.P.H.)

1201 NW 16th Street, 11 GRC, Miami FL 33125

- Michael E. DeBakey VA Medical Center (Olaoluwa Okusaga, M.D.)

2002 Holcombe Blvd, Houston, TX 77030

- Minneapolis VA Health Care System (Maureen Murdoch, M.D., M.P.H.)

One Veterans Drive, Minneapolis, MN 55417

- N. FL/S. GA Veterans Health System (Peruvemba Sriram, M.D.)

1601 SW Archer Road, Gainesville, FL 32608

- Northport VA Medical Center (Shing Shing Yeh, Ph.D., M.D.)

79 Middleville Road, Northport, NY 11768

- Overton Brooks VA Medical Center (Neeraj Tandon, M.D.)

510 East Stoner Ave, Shreveport, LA 71101

- Philadelphia VA Medical Center (Darshana Jhala, M.D.)
3900 Woodland Avenue, Philadelphia, PA 19104
- Phoenix VA Health Care System (Samuel Aguayo, M.D.)
650 E. Indian School Road, Phoenix, AZ 85012
- Portland VA Medical Center (David Cohen, M.D.)
3710 SW U.S. Veterans Hospital Road, Portland, OR 97239
- Providence VA Medical Center (Satish Sharma, M.D.)
830 Chalkstone Avenue, Providence, RI 02908
- Richard Roudebush VA Medical Center (Suthat Liangpunsakul, M.D., M.P.H.)
1481 West 10th Street, Indianapolis, IN 46202
- Salem VA Medical Center (Kris Ann Oursler, M.D.)
1970 Roanoke Blvd, Salem, VA 24153
- San Francisco VA Health Care System (Mary Whooley, M.D.)
4150 Clement Street, San Francisco, CA 94121
- South Texas Veterans Health Care System (Sunil Ahuja, M.D.)
7400 Merton Minter Boulevard, San Antonio, TX 78229
- Southeast Louisiana Veterans Health Care System (Joseph Constans, Ph.D.)
2400 Canal Street, New Orleans, LA 70119
- Southern Arizona VA Health Care System (Paul Meyer, M.D., Ph.D.)
3601 S 6th Avenue, Tucson, AZ 85723
- Sioux Falls VA Health Care System (Jennifer Greco, M.D.)
2501 W 22nd Street, Sioux Falls, SD 57105
- St. Louis VA Health Care System (Michael Rauchman, M.D.)
915 North Grand Blvd, St. Louis, MO 63106
- Syracuse VA Medical Center (Richard Servatius, Ph.D.)
800 Irving Avenue, Syracuse, NY 13210
- VA Eastern Kansas Health Care System (Melinda Gaddy, Ph.D.)
4101 S 4th Street Trafficway, Leavenworth, KS 66048
- VA Greater Los Angeles Health Care System (Agnes Wallbom, M.D., M.S.)
11301 Wilshire Blvd, Los Angeles, CA 90073
- VA Long Beach Healthcare System (Timothy Morgan, M.D.)
5901 East 7th Street Long Beach, CA 90822
- VA Maine Healthcare System (Todd Stapley, D.O.)
1 VA Center, Augusta, ME 04330
- VA New York Harbor Healthcare System (Peter Liang, M.D., M.P.H.)
423 East 23rd Street, New York, NY 10010
- VA Pacific Islands Health Care System (Daryl Fujii, Ph.D.)
459 Patterson Rd, Honolulu, HI 96819
- VA Palo Alto Health Care System (Philip Tsao, Ph.D.)

3801 Miranda Avenue, Palo Alto, CA 94304-1290
- VA Pittsburgh Health Care System (Patrick Strollo, Jr., M.D.)
University Drive, Pittsburgh, PA 15240
- VA Puget Sound Health Care System (Edward Boyko, M.D.)
1660 S. Columbian Way, Seattle, WA 98108-1597
- VA Salt Lake City Health Care System (Jessica Walsh, M.D.)
500 Foothill Drive, Salt Lake City, UT 84148
- VA San Diego Healthcare System (Samir Gupta, M.D., M.S.C.S.)
3350 La Jolla Village Drive, San Diego, CA 92161
- VA Sierra Nevada Health Care System (Mostaqul Huq, Pharm.D., Ph.D.)
975 Kirman Avenue, Reno, NV 89502
- VA Southern Nevada Healthcare System (Joseph Fayad, M.D.)
6900 North Pecos Road, North Las Vegas, NV 89086
- VA Tennessee Valley Healthcare System (Adriana Hung, M.D., M.P.H.)
1310 24th Avenue, South Nashville, TN 37212
- Washington DC VA Medical Center (Jack Lichy, M.D., Ph.D.)
50 Irving St, Washington, D. C. 20422
- W.G. (Bill) Hefner VA Medical Center (Robin Hurley, M.D.)
1601 Brenner Ave, Salisbury, NC 28144
- White River Junction VA Medical Center (Brooks Robey, M.D.)
163 Veterans Drive, White River Junction, VT 05009
- William S. Middleton Memorial Veterans Hospital (Prakash Balasubramanian, M.D.)
2500 Overlook Terrace, Madison, WI 53705

Supplementary Figures

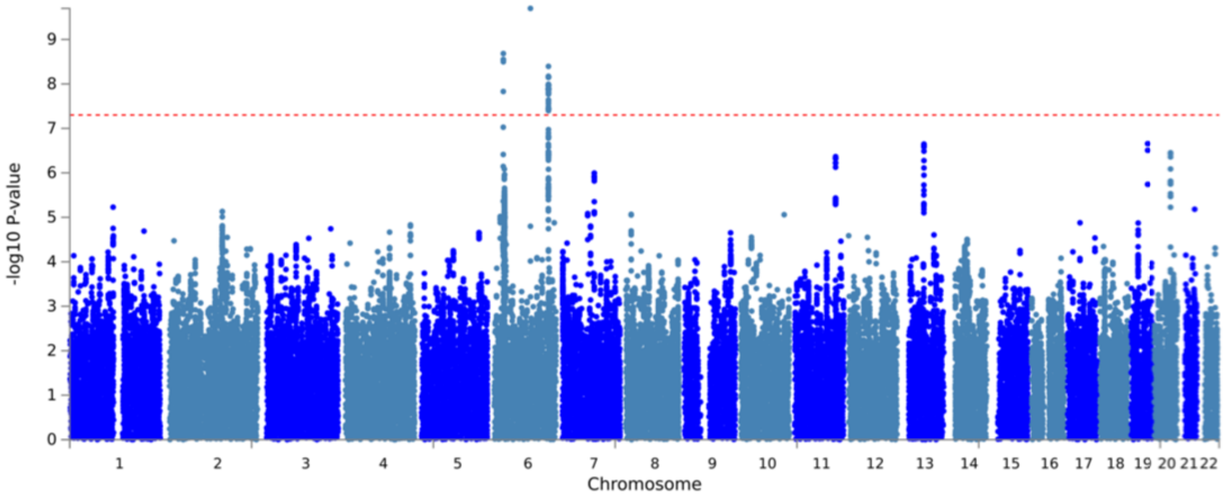


**Supplementary Fig. 1: Multi-ancestry meta analysis GWAS.**
Manhattan plot for the MAMA-MD-BED*BMI GWAS. The *x* axis denotes the chromosome and position of the corresponding SNP. The strength of the SNP-phenotype association is on the *y* axis as the negative $\log_{10}$ of uncorrected two-sided *P* value ($-\log_{10}p$). The dashed gray line represents genome-wide significance ($p = 5.0 \times 10^{-8}$).

**a** Manhattan Plot EUR-ICD-BED

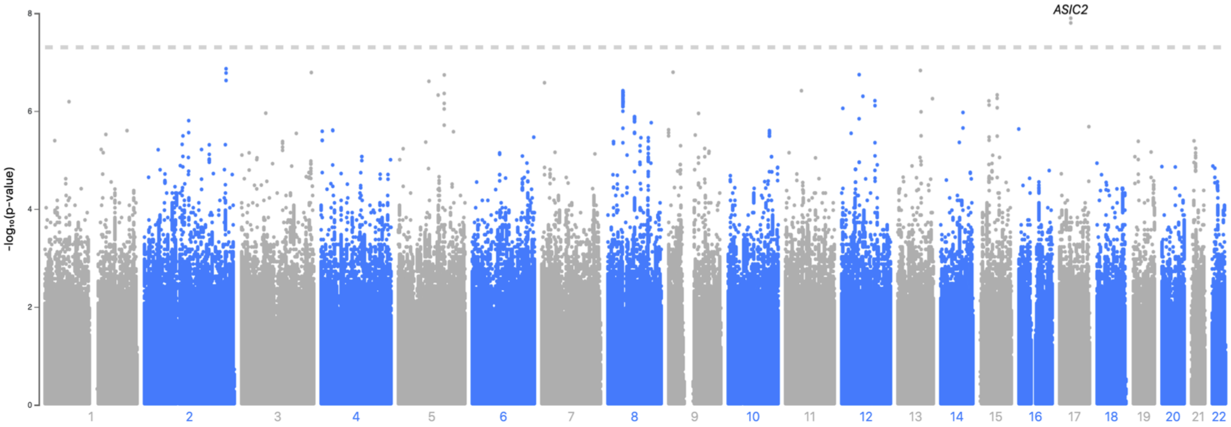


**b** Manhattan Plot EUR-ICD-BED*BMI

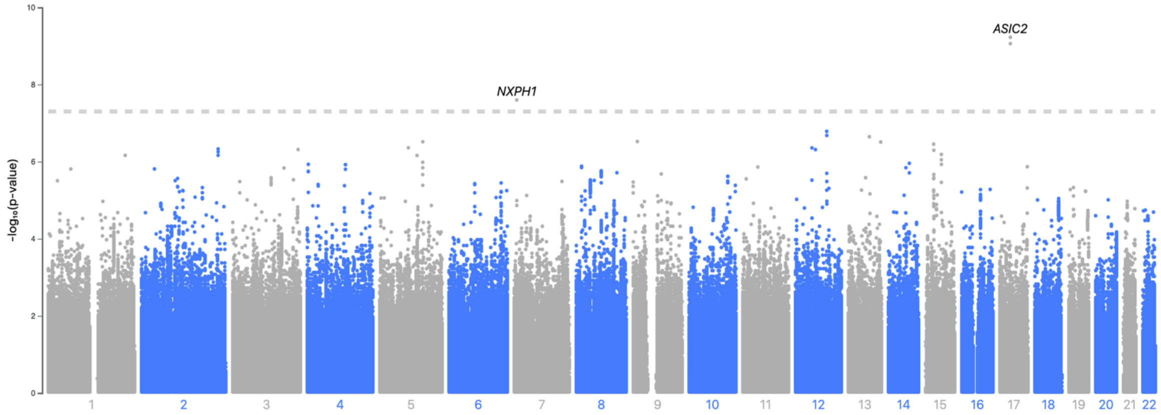


**Supplementary Fig. 2: EUR-ICD-BED and EUR-ICD-BED*BMI GWAS.**

**a-b,** Manhattan plots for the **a,** EUR-ICD-BED and **b,** EUR-ICD-BED*BMI GWAS. The *x* axis denotes the chromosome and position of the corresponding SNP. The strength of the SNP-phenotype association is on the *y* axis as the negative $\log_{10}$ of uncorrected two-sided *P* value ($-\log_{10}p$). The dashed gray line represents genome-wide significance ($p = 5.0\times10^{-8}$).

**S3a**

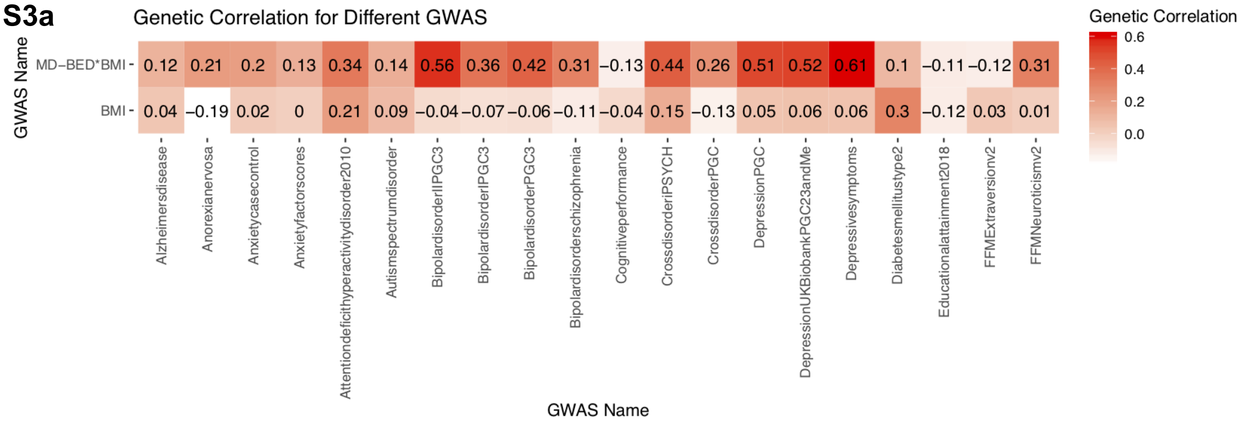


**S3b**

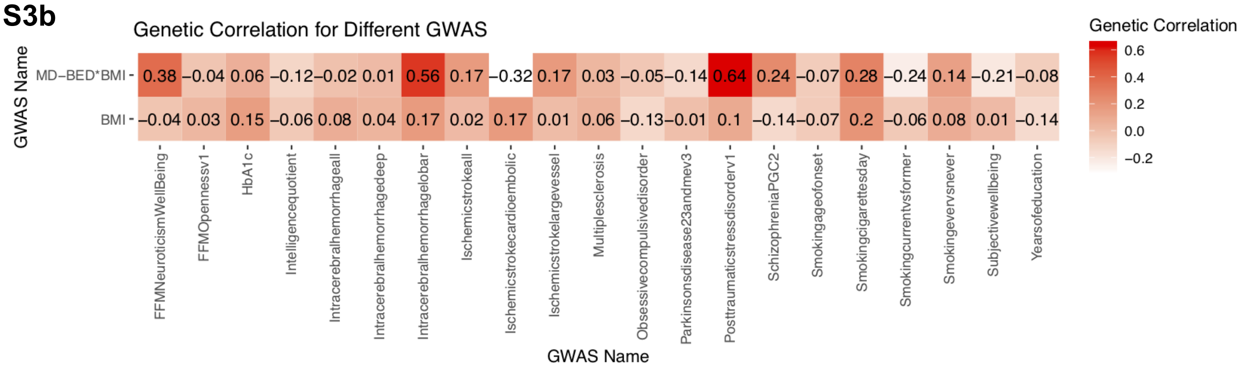


**Supplementary Fig. 3: Genetic correlation between BED, BMI and other traits.**

**S3a-b,** Correlation matrices where the *y* axis denotes primary GWAS and the *x* axis lists comparison trait GWAS. Cells include the genetic correlation between the two GWAS, with color corresponding to the strength of the genetic correlation.

**S3c**

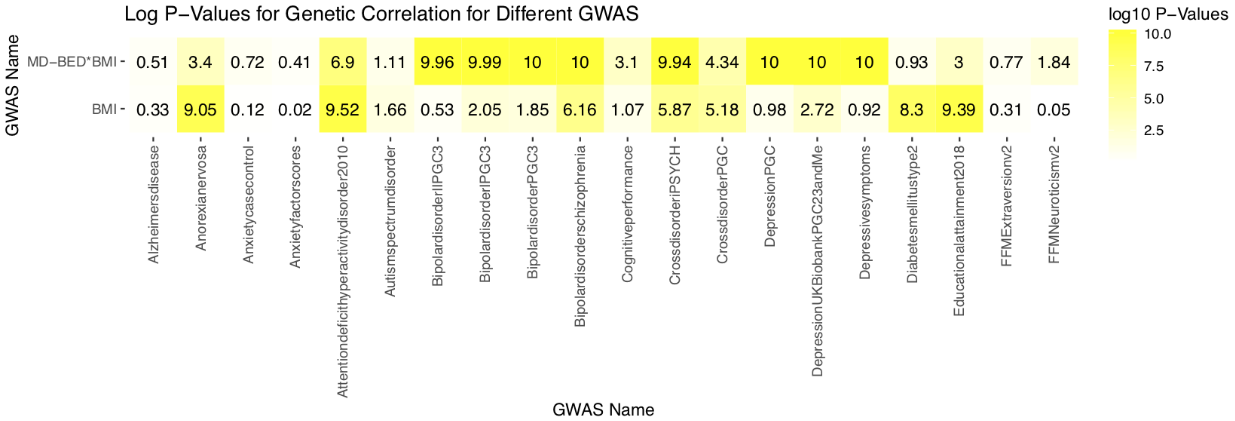


**S3d**

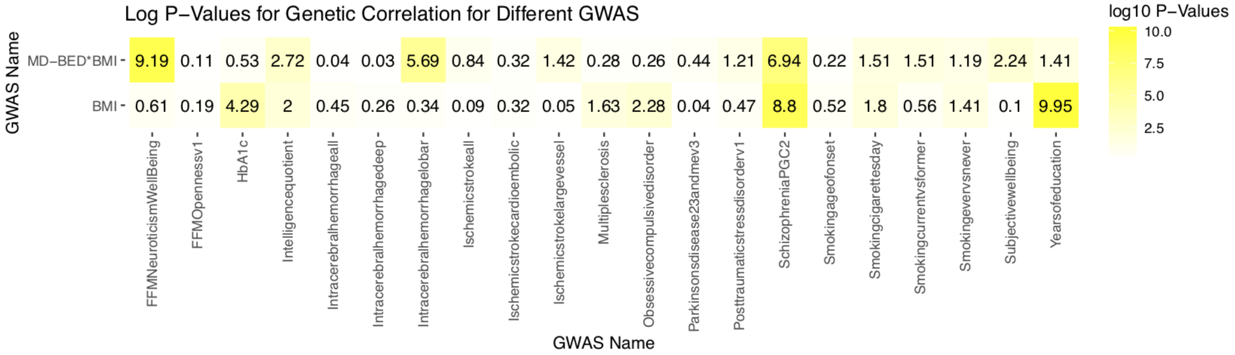


**Supplementary Fig. 3 (continued): Genetic correlation between BED, BMI and other traits. S3c-d,** *P* value matrices where the *y* axis denotes different BED related phenotypes and the *x* axis lists comparison trait GWAS. Cells include the negative $\log_{10}$ of uncorrected two-sided *P* value colored by *P* value magnitude ($-\log_{10}p$).

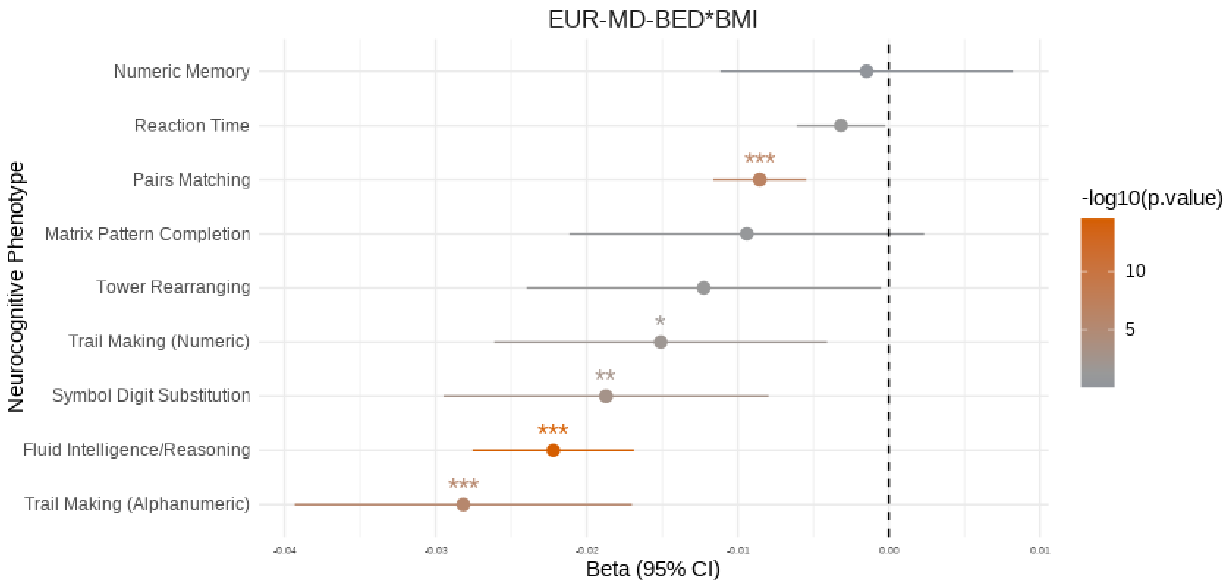


**Supplementary Fig. 4: Neurocognitive Traits.**
EUR-MD-BED*BMI PRS association with neurocognitive traits measured in the UKBB. Neurocognitive measures from the UKBB are shown on the *y* axis. Beta is shown on the *x* axis. The dashed black line is at 0 on the *x* axis. Confidence intervals are two-sided 95% confidence intervals (CI). The significance of the association is shown as negative $\log_{10}$ of adjusted two-sided *P* value ($-\log_{10}$p) through color of the point. *$p < 0.05$. **$p < 0.01$. ***$p < 0.001$.

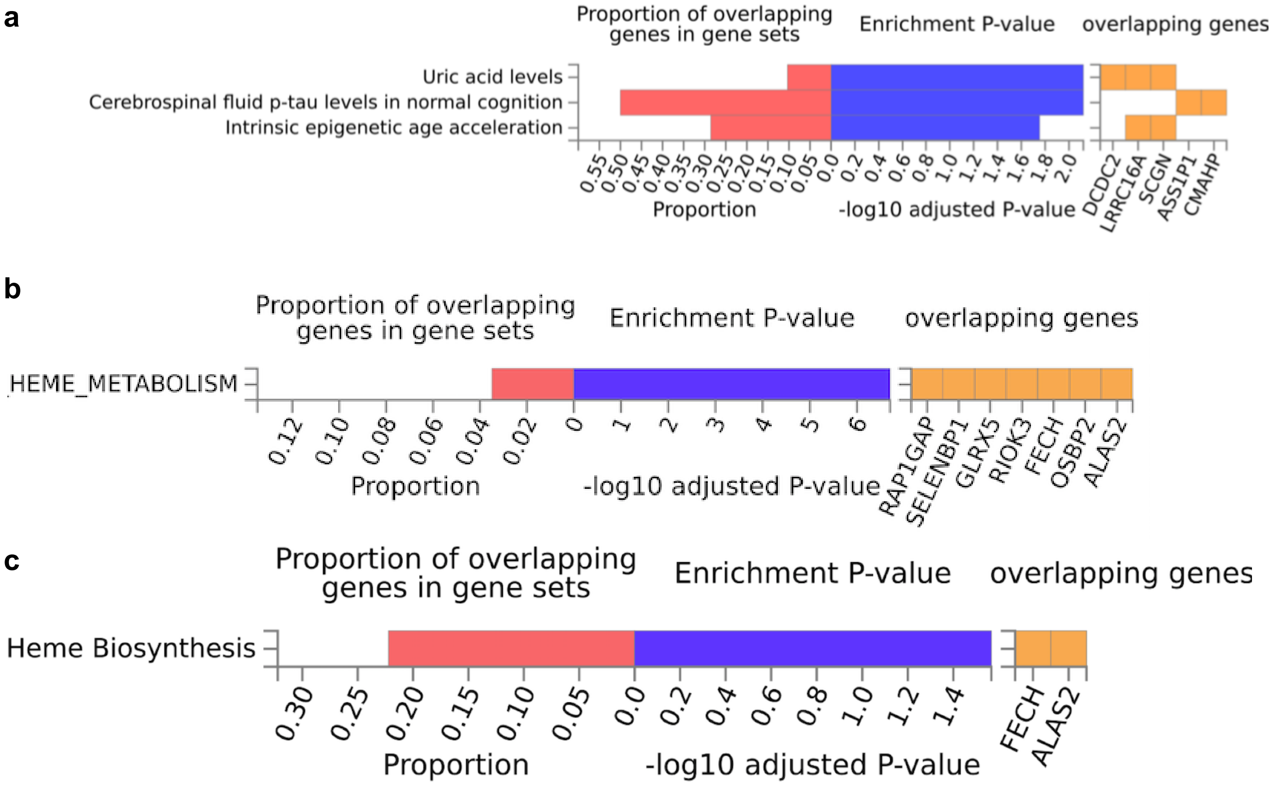


**Supplementary Fig. 5: FUMA gene set enrichment for MD-BED*BMI.**

**a-c,** The *y* axes denotes phenotypes that are enriched for genes similar to the MD-BED*BMI GWAS. The leftmost plots show the proportion of genes in the gene set that share enrichment. The middle plots show the negative $\log_{10}$ of adjusted two-sided *P* value ($-\log_{10}p$). The rightmost plots highlight individual genes that are enriched in both phenotypes.

**S6a**

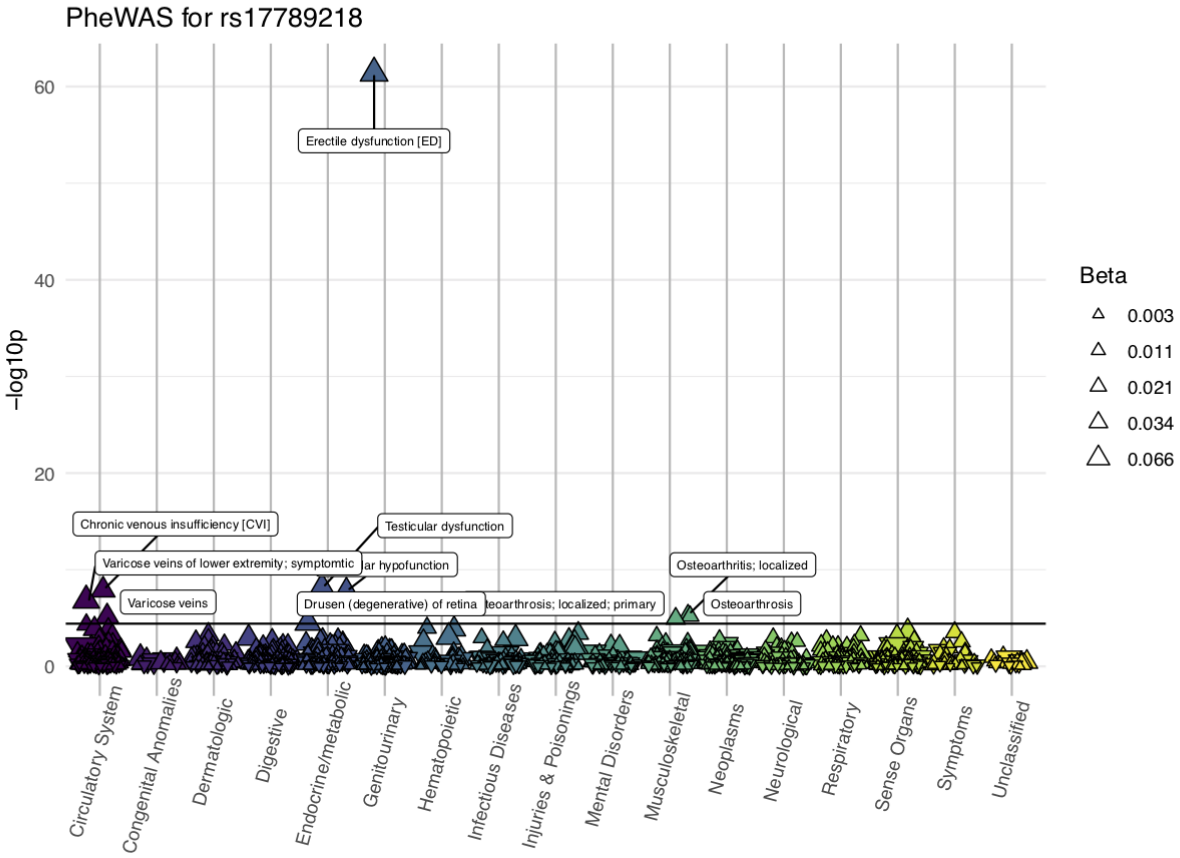


**Supplementary Fig. 6: PheWAS for genome-wide significant loci.**
Scatter plots for the three lead SNPs from genome-wide significant loci from the FEMA-MD-BED*BMI GWAS (**S6a**: rs17789218). The categories of phenotypes are listed on the *x* axis. The transformed negative $\log_{10}$ $Q$ value is represented on the *y* axis. The black line represents significance. The direction of the triangle indicates whether the regression coefficient is positive (up) or negative (down) and the size of the triangle indicates the magnitude of the regression coefficient.

**S6b**

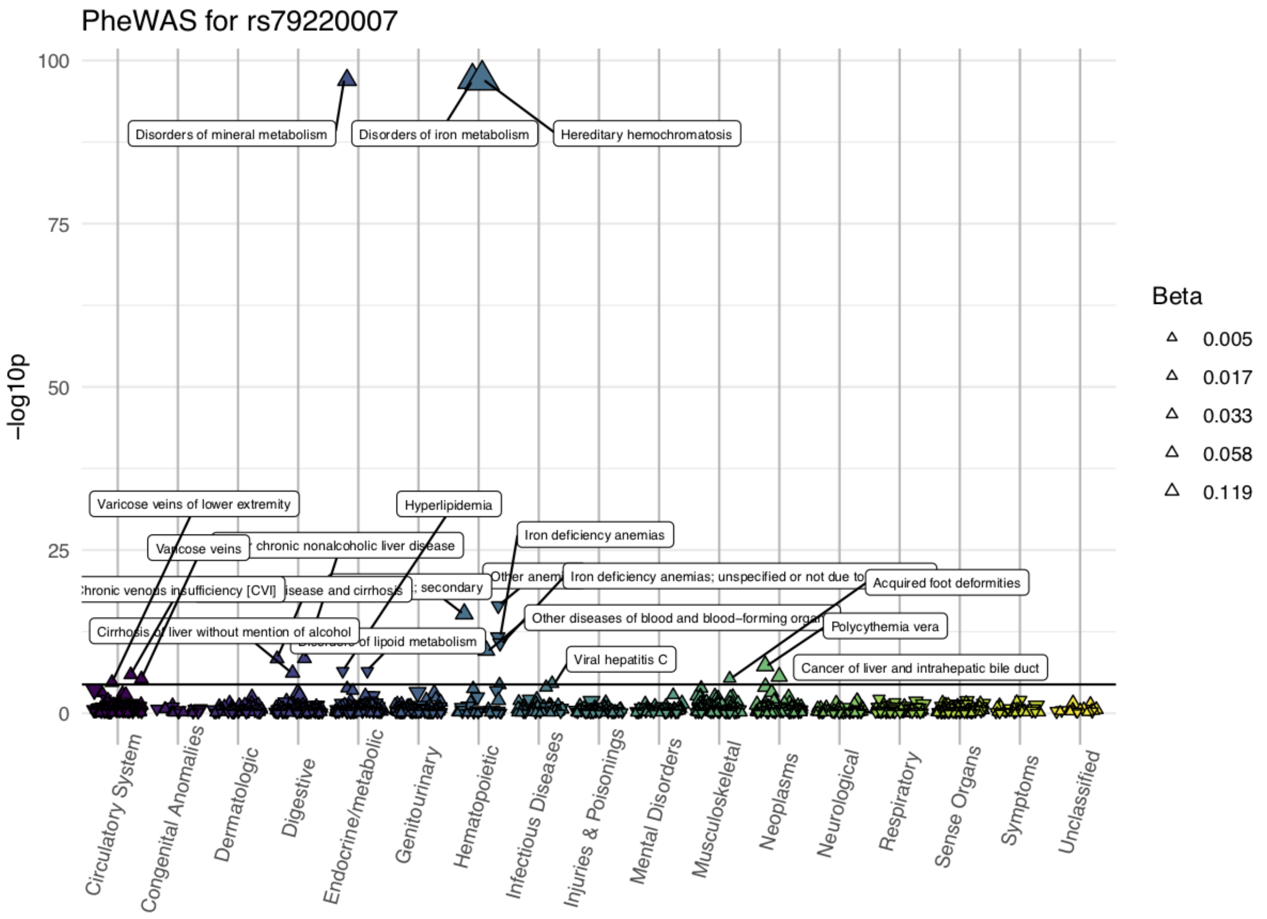


**Supplementary Fig. 6 (continued): PheWAS for genome-wide significant loci.**

Scatter plots for the three lead SNPs from genome-wide significant loci from the FEMA-MD-BED*BMI GWAS (**S6b**: rs79220007). The categories of phenotypes are listed on the *x* axis. The transformed negative $\log_{10}$ $Q$ value is represented on the *y* axis. The black line represents significance. The direction of the triangle indicates whether the regression coefficient is positive (up) or negative (down) and the size of the triangle indicates the magnitude of the regression coefficient.

**S6c**

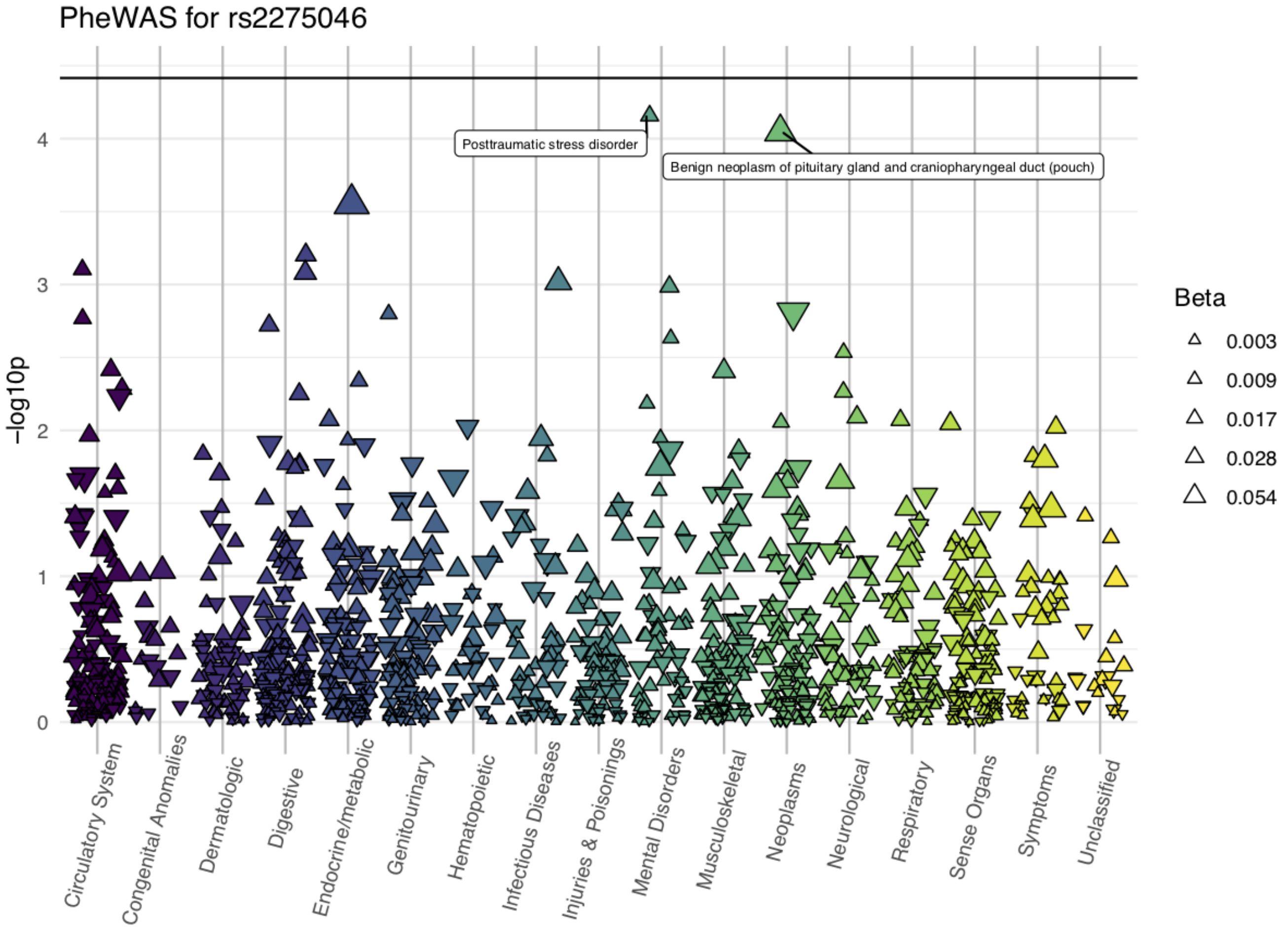


**Supplementary Fig. 6 (continued): PheWAS for genome-wide significant loci.**

Scatter plots for the three lead SNPs from genome-wide significant loci from the FEMA-MD-BED*BMI GWAS (**S6c**: rs2275046). The categories of phenotypes are listed on the *x* axis. The transformed negative $\log_{10}$ *Q* value is represented on the *y* axis. The black line represents significance. The direction of the triangle indicates whether the regression coefficient is positive (up) or negative (down) and the size of the triangle indicates the magnitude of the regression coefficient.

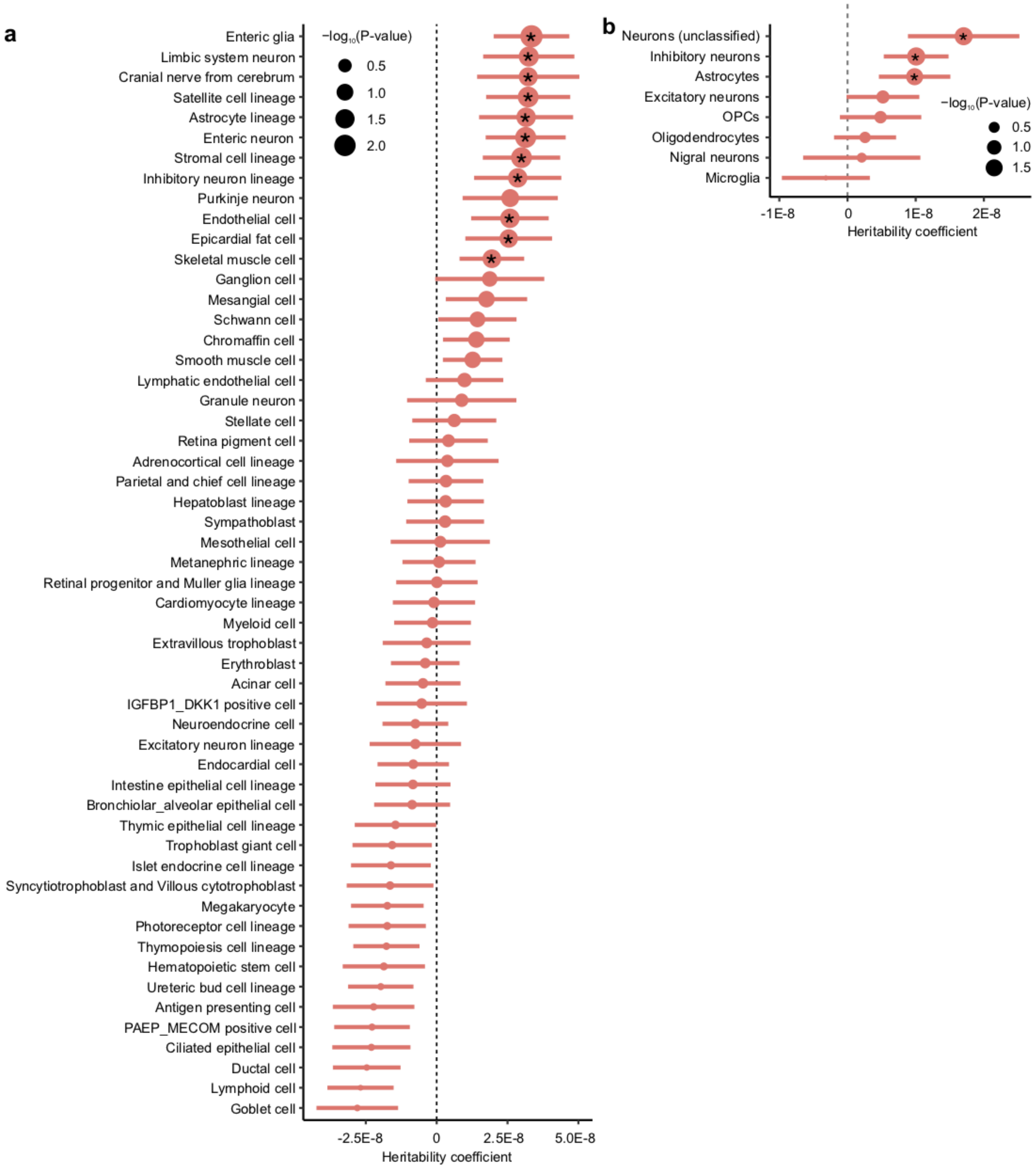


**Supplementary Fig. 7: Cell-type enrichment.**

**a-b,** Enrichment of BED risk variants in genomic regions of open chromatin in **a,** a variety of fetal cell types and **b,** brain-specific adult cell types. A positive coefficient signifies enrichment in heritability. Dot size reflects negative $\log_{10}$ of uncorrected two-sided $P$ value ($-\log_{10}$p) of the LD-score regression. *Nominally significant, $p < 0.05$.

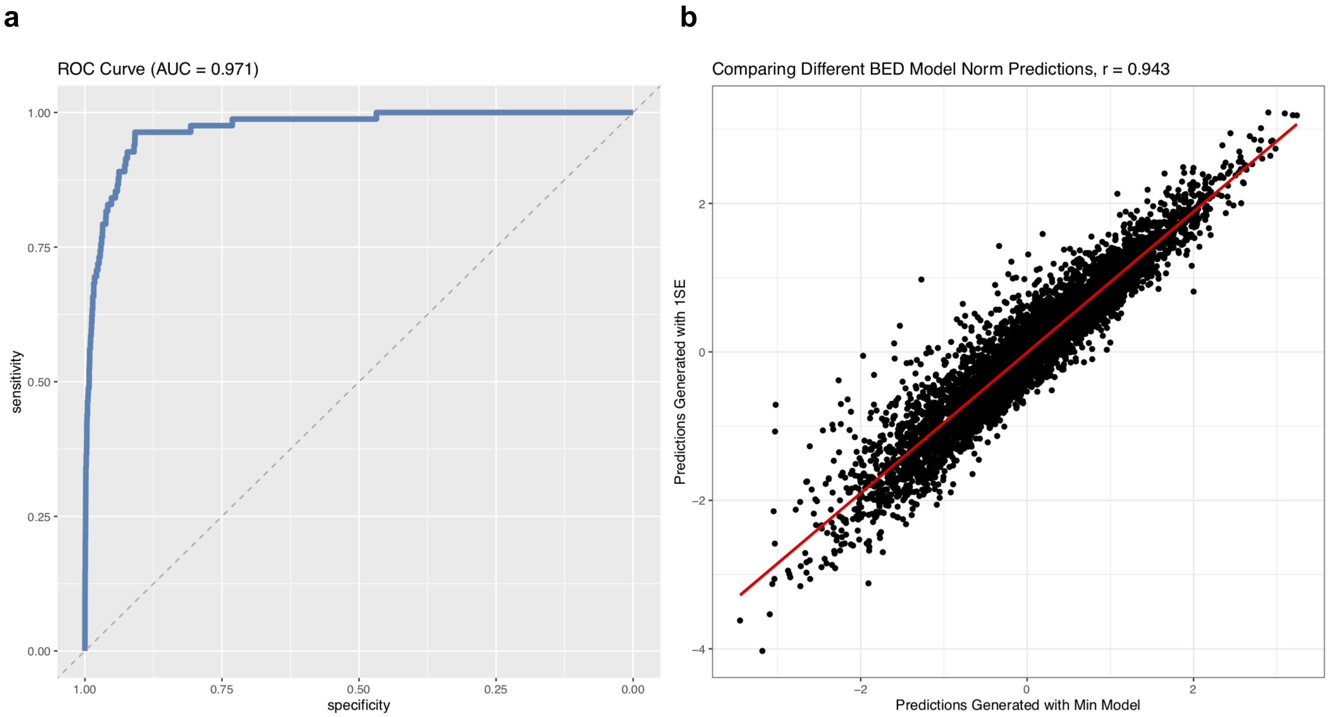


**Supplementary Fig. 8: Model Selection for Predicting BED in the MVP.**
**a,** Receiver operating characteristic (ROC) curve (dark blue line) for predicting BED in a stratified test set consisting of 10% of the data. The *x* axis shows the false positive rate (specificity in reverse direction) and the *y* axis shows the true positive rate (sensitivity). The dashed gray line represents chance performance. The area under the curve (AUC) is 0.97. **b,** Scatter plot comparing the inverse-rank normal transformed predictions from the model with parameters that minimizes binomial deviance through 10-fold cross validation (Min) and the simplest model that falls within one standard deviation of the optimal binomial deviance (1SE).

### Supplementary Results

#### Uric Acid Metabolism

To determine the relationship between uric acid metabolism and BED, we first examined the association of our EUR-MD-BED*BMI and EUR-BMI GWAS PRS with baseline urate levels from the UKBB (Table S7). Without adjusting for BMI, urate levels were strongly correlated with EUR-BMI ($\beta$ = 0.098, $p$ = $1.50\times10^{-206}$) and were negatively correlated with EUR-MD-BED*BMI ($\beta$ = -0.025, $p$ = $1.21\times10^{-15}$). However, after adjusting for BMI, the EUR-BMI and EUR-MD-BED*BMI PRS were both negatively correlated with urate (EUR-BMI: $\beta$ = -0.063, $p$ = $3.73\times10^{-96}$; EUR-MD-BED*BMI: $\beta$ = -0.022, $p$ = $1.22\times10^{-13}$). We also examined the relationship between our EUR-MD-BED*BMI and EUR-BMI GWAS PRS and gout in the UKBB. Similar to urate levels, we found a strong positive correlation between gout and the EUR-BMI PRS ($\beta$ = 0.09, $p$ = $7.17\times10^{-14}$) as well as a negative correlation with EUR-MD-BED*BMI ($\beta$ = -0.037, $p$ = $1.72\times10^{-3}$) when our model did not account for BMI. After adjusting gout risk for BMI, both the EUR-BMI and EUR-MD-BED*BMI PRS were negatively correlated with gout (EUR-BMI: $\beta$ = -0.057, $p$ =$4.30\times10^{-6}$; EUR-MD-BED*BMI: $\beta$ = -0.034, $p$ = $4.88\times10^{-3}$). We then leveraged GSMR to assess causality between urate levels and the EUR-MD-BED*BMI phenotype and observed a weak negative relationship between the two phenotypes ($\beta$ = -0.007, $p$ = .061).

While these PRS association studies found negative correlations with gout and urate levels, obese individuals with BED have been found to have higher urate levels compared to obese controls[1]. Numerous factors, including sex, renal function, diuretic use, diet and BMI impact urate levels and demographic differences between these variables within the MVP and UKBB as well as BED-specific environmental confounds may have impacted our findings[2,3]. Whether and how uric acid metabolism interplays with BED remains unanswered.

#### EUR-ICD-BED and EUR-ICD-BED GWAS

The EUR-ICD-BED GWAS found a single locus reaching genome-wide significance on chromosome 17 at *ASIC2* (Supplementary Table 5). This locus survived BMI-adjustment in the EUR-ICD-BED*BMI GWAS, which identified an additional genome-wide significant locus on chromosome 7 at *NXPH1* (Supplementary Table 5). We used MAGMA[4] to identify protein coding genes associated with the aforementioned phenotypes and found a significant association between *KAT6A* and the EUR-ICD-BED phenotype ($p$ < $6.43\times10^{-7}$). These results should be interpreted cautiously as heritability of the EUR-ICD-BED GWAS was nominally significant ($h^2$ = 22.3-29.5%, $p$ = 0.05) and heritability of the EUR-ICD-BED*BMI GWAS was not significant ($h^2$ = 16.9-22.4%, $p$ = 0.11).

#### Cell type enrichment of risk variants

To identify cell types in which the genetic drivers of BED are likely to have an effect, we compared our EUR-MD-BED*BMI GWAS to two chromatin accessibility atlases. First, we used a comprehensive atlas of human fetal development to take advantage of the diversity of tissue types available for comparison[5]. While no individual cell types were significantly associated with BED at the false discovery rate of 0.05, we saw nominally significant enrichment across neural lineages including from limbic system neurons, the cranial nerve, inhibitory neurons, enteric neurons, enteric glia, astrocytes, and from stromal and satellite cell lineages (Supplementary Fig. 7a). A similar enrichment of non-excitatory neurons and astrocytes was recapitulated using an adult brain atlas[6] (Supplementary Fig. 7b).
